# Profiles of Social Distance Compliance: Psychological and Situational Predictors of Risky Behavior during COVID-19

**DOI:** 10.1101/2020.06.04.20122754

**Authors:** Michael Robert Haupt, Staci Meredith Weiss, Michelle Chiu, Raphael Cuomo, Jason M. Chein, Tim Mackey

**Affiliations:** Department of Cognitive Science, University of California San Diego, 9500 Gilman Dr, La Jolla, CA, 92093, USA; Department of Psychology, Temple University, 1701 N. 13th Street, Philadelphia, PA 19122, USA; Department of Psychology, University of Cambridge, Downing St, Cambridge CB2 3EB, United Kingdom; Department of Anesthesiology and Division of Infectious Diseases and Global Public Health, University of California San Diego School of Medicine, 9500 Gilman Dr, La Jolla, CA, 92093, USA

## Abstract

The purpose of this study was to explore the factors underlying variability in compliance with CDC guidelines in response to the novel coronavirus, or COVID-19. To do this, we examined the frequency of once ordinary, but newly risky behavior (as deemed by CDC guidelines) in a sample of 482 MTurkers. We ran analyses probing the situational and dispositional variables that predicted variance in risky behavior using data-driven and hypothesis-generated approaches. We found situational and dispositional variables contributed unique variance to risky behavior, controlling for variability accounted for by demographic factors. More frequent report of risky activity was associated with *higher* extraversion, need for cognitive closure, behavior activation, and perceived resource scarcity; in contrast, more frequent report of risky activity was associated with *less* empathy and living space access, as well as *younger* age. To break down these findings, we used a cluster analysis to profile individuals, using only situational and dispositional variables belonging to seven clusters. Combined with testing differences in risk taking by cluster identity, we suggest this profile approach might allow consideration of multi-faceted attributes that influence adherence with public health guidance in the context of health emergencies like the COVID-19 pandemic.

## Background on COVID-19

Since March 2020, the lives of citizens across the United States (US) and around the globe have been upended by the emergence of the COVID-19 pandemic. For some countries, this change has occurred even sooner, with China reporting cases of the virus as early as November 2019^1^. On March 23, the Centers for Disease Control and Prevention (CDC) distributed the first set of guidelines for how individuals can mitigate their ‘risk’ for Coronavirus infection and contagion. This advice introduced “social distance” into public discourse, imploring individuals to minimize physical proximity to others outside of their household by maintaining 6 feet of distance with others when interacting, outdoors or in public areas. While these measures differ based on jurisdiction (including variation at the local, city, municipal, state, provincial, and country level), in the United States, many countries have closed down public areas to avoid mass gatherings, and many local businesses and restaurants have had to close or rely on online commerce options in order to continue operations^2, 3^.

On April 2, the CDC updated their recommendations, advising the use of face coverings or masks in situations “at risk” for violations of social distance, and urging avoidance of unnecessary exposure at visits to businesses or public spaces^4^. The tone of public health authorities had shifted from suggesting augmented activity to imploring minimized activity, such that recently routine activities were newly associated with confronting “risk” of harm to oneself or others. In the absence of a federal mandate, by April 10th more than 95% of the American population was under advisement to minimize their activity and risk for infection, as state governments and local municipalities enacted ‘Stay at Home’ recommendations or ‘Shelter-in-place’ orders^5^. These recommendations advised citizens to stay indoors and only venture outside of one’s residence for ‘essential’ errands. Additionally, many advertising campaigns promoted social distancing as a form of social and personal responsibility such as the statewide, multimedia “Stay Safe, Stay Home” campaign^6^ and a campaign lead by Healthcare leaders (including a former U.S. Surgeon General) in which they ask that people to “stay at home as much as possible” and “avoid all crowds”^7^. However, despite the cooperation by government, business and authorities to promote (and in some cases, enforce) social distancing, many members of the public show signs of restlessness towards stay at home orders, even as confirmed case counts and the mortality rates do not experience sharp declines^8^.

Among Americans, there is variability in the impact of COVID-19 on disruption of employment, routine and livelihood, and adherence to CDC guidelines is likely to be dependent on individual circumstances and situational factors. Situational factors are understood as employment, living space and resource access, which may face disruption related to COVID-19. For example, the ability to work remotely is related to education, digital access, child and/or elder care responsibilities, physical infrastructure and countless other factors, which in turn are highly stratified by socioeconomic and ethnic demographics. Thus, the *ability* to comply with social distance guidelines may vary based upon demographic differences.

Recent studies show evidence, for instance, that Black/African Americans are being disproportionately infected by COVID-19^9^, while data from New York City show higher COVID-19 related death rates for both Black/African and Hispanic/Latino persons^10^. People categorized as essential workers in Health, Food/Agriculture, and Infrastructure industrial sectors receive some financial security by remaining employed, but also put themselves at higher risk of contracting the virus due to increased exposure^11^. Additionally, over a quarter of private sector workers in the United States do not receive paid sick leave (including over 30% of workers in the South and Midwest), which could cause further spread of the disease that disproportionately impacts certain at-risk populations^12, 13^. Hence, social determinants of health, such as poverty, ethnicity, employment status, healthcare access, and other known factors are likely exacerbated and accentuated for already vulnerable populations in the face of COVID-19.

## Reconceptualizing Risky Behavior

The saturation of news coverage relating to death, injury and illness has made the looming threat of COVID-19 ubiquitous, such that activities that were recently routine (and perceived necessary) have been identified as conferring ‘risk’ or to oneself or others^9^, with potentially deadly consequences. In response to COVID-19, reports of heightened anxiety, depression, and posttraumatic stress have been associated with variability in personality^14^. Early reports from cases in China^15^, as well as studies under review examining Italian and Brazilian populations, have indicated that a range of psychological, situational and demographic factors impact stress related to COVID-19, coping with social distance and/or limited social contact, and even COVID-19 health outcomes^16, 17^. Thus far, these studies have focused on attitudes towards COVID-19, stay-at-home guidance and social distance measures, rather than incremental changes and rapid shifts in beliefs and behavior related to compliance with public health guidance. Compounded by the role of a public authority compelling individuals to modify their behavior and habits, it may be adaptive for individuals to alter their beliefs about the consequences or risk involved in neglecting public health guidance. Prior studies have tracked public sentiment during crises such as the H1N1 pandemic^18^ and linked public sentiment during global health and natural disasters (including the 2003 SARS outbreak) with changes in personal risk perception, risk-to-benefit evaluations of behavior and health-related compliance behavior^19^.

Interestingly, salient threats to an individual’s mortality have been associated with both greater risk aversion^20, 21^ and greater risk-inclination^22^. In the first account consistent with Terror Management Theory, under threat, self-protective tendencies reign and behavior reflects extreme risk aversion^23^; in the second account consistent with Mortality Salience, self-destructive tendencies reign, control of behavior is depleted and individuals may evaluate the risk for harm as lower and/or less likely to affect them^22^. Variability in risk taking and attitudes in response to threat are partially accounted for by individual differences in age, as well as disposition and personality^24^. Greater risk taking is also associated with lower inhibition, higher sensation-seeking, and lesser need for cognitive closure^25, 26, 27^

Medical conditions, disability diagnosis and physical injury can disrupt routines and alter perceptions of risk; reciprocally, personality traits related to risk-taking propensity, such as inhibition, sensation-seeking, extraversion and poor self-regulation abilities have all been associated with a higher incidence of risky behaviors that may lead to adverse health outcomes^28, 29^. For example, in adolescents with childhood cancer experience and adults undergoing chemotherapy, vigilance against health risks is generalized, impacting risk sensitivity and evaluation more broadly^30^. Risk taking in patients with ‘bedrest’ or ‘homebound’ recommendations due to their vulnerable condition or compromised immune systems might provide an interesting parallel to individuals responses to the COVID-19 stay at home and self-isolation orders^30^. The relevance of these psychological factors to COVID-19 risk taking and social distance compliance is uncertain as people likely react differently to the multiple, interacting aspects of the public health guidance such as the salient mortality threat and heightened uncertainty, the mandate to avoid physical proximity to other people and the imperative to enact precautions which prevent contracting and spreading infection.

## Influences on Risk Taking and Compliance with Public Health Guidance

Adjusting quarantine guidelines based on a deeper understanding of psychological and situational factors can potentially improve public health outcomes, especially in the face of increased resistance to compliance under current guidelines that continues to grow after just a few weeks since quarantine restrictions have been implemented in the US. During the middle of April 2020, “Liberate” protests were organized around city halls in states such as Minnesota, Michigan, and Virginia where protesters demanded an end to lockdown restrictions^31^. Even though polls show that the majority of Americans do not support the Liberate protests^32^, there may still be widespread variability in compliance with public health guidelines. One week after the protests, tens of thousands of people packed beaches in Southern California, violating social distancing guidelines^33^. Concomitantly, smartphone movement data revealed a significant decline in social distance adherence on April 14th, after three weeks of data consistent with compliance^34^. Hence, though most citizens do not actively protest or resent stay at home guidelines^32^, situational, dispositional and demographic factors may explain why individuals struggle to comply with stay-at-home orders and social distancing requirements. From a public health standpoint, ‘quarantine fatigue’, inequities in the ability to comply with public health guidance, and Liberate protests represent breaks in the firewall of COVID-19 public health response and contagion control, serving as a potential threat to crucial gains made in “flattening” the COVID-19 epidemiological curve.

While the impact of the COVID-19 pandemic is widespread, it appears that people are experiencing different contextual challenges which could influence compliance with social distancing measures. In this study we examined the association of variability in psychological *dispositions* (personality, need for cognitive closure, empathy, and sensation-seeking) and *situational factors* (age, employment, living space access and perceived resource scarcity) in relation to a newly developed measure of COVID-19 *risk taking* (as an index of compliance with social distance measures and stay-at-home guidelines), along with *demographic* features. In order to further unpack the individual differences associated with COVID-19 risk taking, a *k*-means clustering algorithm was deployed to identify distinct *trait and situational profiles*. Using trait and situational variables as inputs, we found variability among clusters in the risky behavior which might indicate non-compliance with the stay-at-home measures. In addition, we found cluster differences in data-driven group classifications of ‘risk propensity.’ We further assessed variability by cluster in risky behavior and risk perception, now that engaging in once-mundane activity has the potential to inflict harm to oneself or others. These multivariate approaches afford high-level generalizations that facilitate how researchers and public health officials conceptualize ‘types’ of individuals within the US population, with the goal of providing tailored, realistic, effective public health messaging to mitigate the spread of COVID-19.

## Methods

Our survey was developed and piloted in a small sample of undergraduates for distribution via Qualtrics. It used a range of existing psychological measures and batteries, as well as our own and others newly-developed items which were created to interrogate aspects specific to COVID-19. Participants were consented and compensated at a rate of $7/hour for completing the 30-minute survey, referred from the Amazon Mechanical Turk (MTurk) database of contractors.

### Participants

A total of 514 participants were recruited from Amazon Mechanical Turk (MTurk) with the aim to have a sample size approaching n = 500 (aiming for a power of .80 for even smaller effect sizes), after filtering participants who did not pass data quality checks^35^. Recruitment occurred between April 30^th^ and May 2^nd^ following the first full month of quarantine within the United States. Final sample size was n = 482 after removing poor quality and/or incomplete responses. Data quality checks included: speed outliers and/or incomplete responses (determined using Mahalnobi’s distance, M = 32.5 minutes to complete, n = 9); response to the question “Estimate the Date you first modified your behavior due to the Coronavirus” (to identify malingerers, n = 20); and duplicate MID or completion codes (n = 3).

Our sample demographics roughly reflected the ethnic breakdown of the American public according to 2014–2018 Census data, with a significant underrepresentation of individuals identifying as ethnically Hispanic^36^. Age within the sample ranged from 18 to 73 with an average age of 37.12 (SD = 11.33) and 59% of participants identified as men. Additionally, 71.2% of participants were White, 19.7% Black, 6.0% Asian, and 7.1% Hispanic. See Table A2 in Appendix for more demographic information about the full sample. Ethics approval for this study was received and granted by Temple University IRB. MTurk participants were compensated based on standard survey-taking rates on the platform.

### Self-report measures

*COVID Risk Taking Inventory (CRI)*. A ten-item questionnaire was developed, adapted from the structure and format based on the Benthin Risk Perception Scale^37^, to assess a set of activities (e.g. attending a gathering of more than 5 people) that under the conditions of the COVID-19 pandemic are considered “risky”. Items were developed to target the discrepancies in activities identified as ‘risky’ per CDC guidelines in April, as opposed to those released originally on March 23. Each activity item was followed by four questions that asked about the frequency of engagement since the estimated date when respondents first began modifying their behavior due to COVID, a risk-benefit comparison, and risk propensity toward the self or others. The Cronbach’s alpha for the full scale was above 0.90 for all the ten items, across each of the four sub-questions: risk behavior frequency, cost-benefit evaluation, risk to self, and risk to other. This measure served as our primary outcome variable. Factor analysis was used to identify which behaviors were high-risk, low-risk and essential (travel-related items were not included in further analysis). For a full detailed index of the items and questions used, see Supplementary Information.

### Situational Factors

In assessing situational factors that may influence self-reported behavior, participants were asked to respond to questions relevant to Living Space Access (“How many rooms within your current residence do you feel comfortable relaxing or spending time in that are not your bedroom? This can also include outdoor spaces that are on your property”) and Perceived Scarcity of resources (“It has been difficult for me to get needed resources (food, toilet paper) due to the Coronavirus”), the latter of which was rated on a 7-point Likert scale ranging from 1 (“strongly disagree”) to 7 (“strongly agree”).

### Big Five Personality Inventory

A 23-item questionnaire based on the Big Five Inventory^38, 39, 40^ was used to evaluate participants across the Big Five personality dimensions (extraversion, agreeableness, conscientiousness, neuroticism, openness).

### Empathy

Empathy was assessed by having participants respond to the 7-item Perspective Taking subscale taken directly from the Interpersonal Reactivity Index^41^, with each item rated on a 5-point Likert scale ranging from 1 (“does not describe me well”) to 5 (“describes me well”).

### Need for Cognitive Closure Scale

All participants also completed a 14-item version of the Need for Cognitive Closure (NFCC) Scale^42^. The adapted questionnaire aimed to measure cognitive closure through 5 subscales: order (e.g. “I prefer to have a clear and structured mode of life”), predictability (e.g. “I dislike situations that are unpredictable”), ambiguity (e.g. “I feel uncomfortable when I don’t understand the reason why an event occurred in my life”), decisiveness (e.g. “When I have made a decision, I feel relieved”), and closed-mindedness (e.g. “I feel irritated when one person disagrees with what everyone else in a group believes”). The scales were rated on a 6-point Likert scale ranging from 1 (“strongly disagree”) to 6 (“strongly agree”).

### Behavioral Inhibition/Behavioral Activation Scales

The Behavioral Inhibition/Behavioral Activation Scales^43^ index the two motivational systems of Behavioral Inhibition and Behavioral Activation^44, 45^. The BIS scale includes a subscale measuring sensitivity to aversive motivation (e.g. “criticism or scolding hurts me quite a bit”). The BAS scale measures sensitivity to the mechanism underlying appetitive motivation by using three subscales, namely: drive (e.g. “When good things happen to me, it affects me strongly”), fun-seeking (e.g. “I often act on the spur of the moment”), and reward responsiveness (e.g. “When I get something I want, I feel excited and energized”).

### Analysis

Analyses were conducted in R and SPSS statistical software. First, we identified which of the activities (marked as risky according to the CDC) were considered to be ‘*risky behavior*’ by the participants and then created a mean index using only the subset of items rated as above average on ‘riskier than the benefit’ by our sample. In order to investigate the variables that contribute most to risk-taking behavior, we implemented a Shapley Value regression, which assesses the relative importance of all the independent variables within a model by first computing all possible combinations of the independent variables, and then determining how much each variable contributes to the total R^2^ of the model (see Budescu 1993 for a more detailed description)^46^. For our analysis, a Shapley regression predicting variability in ‘risky behavior’ was run on all dispositional and situational variables. Next, we ran a series of multiple regressions, which aimed to examine whether the dispositional and situational variables that contributed significantly to variance in risky behavior, while also controlling for variance accounted for by demographic factors. We then parsed out whether this relationship held true for all relevant types of activities, which were classified as ‘essential’, ‘low-risk’, ‘high-risk’ and ‘travel’ using a factor analysis on the 10-item COVID Risk-taking Inventory (CRI). To facilitate the interpretation of these regressions and understand why some individuals differed in their COVID risk taking, we used cluster analyses to identify personality traits and contextual factors relevant to compliance with CDC guidelines.

### *k*-means Clustering

Groups for the analysis were created using *k*-means clustering using variables related to psychological and situational circumstances. The final input variables used in the model in this paper are: introversion scores measured by the BFI, sensation seeking scores measured by BIS/BAS, perspective-taking empathy scores from the IRI scale, age, living space access (whether or not the participant lives in a residence with more than 2 common spaces), and perceived scarcity (how much participants agree with the statement “it has been difficult for me to get needed resources (food, toilet paper) due to the Coronavirus” (1 Strong disagree to 7 Strongly agree). Since variables with larger values contribute more to the distance measure in k-means clustering than variables with smaller values^47^ we converted the psychometric scales into binary variables using the sample median score pertaining to each trait, such that participants with scores below the median are classified as low level and participants with scores above the median are classified as high level. By converting the variables into binaries, we prevented scales with larger ranges from overcontributing to the model. Additionally, Living Space Access (number of common spaces) was re-coded into a binary variable from the original 5-point scale, wherein responses were split into two groups, those who responded between 0 and 3 and those who responded 3 or above. We also recoded Age into a 4-point scale based on quartile scores of the original continuous numeric variable.

The final cluster model is the result of an iterative process, which tested different combinations of input variables and group number. In total, 20 models were created and tested during analysis. A model was considered viable if it met the following criteria: Each group within the cluster model must show differentiation from one another, every input variable must have unique relationships with each group in the model (i.e., no two input variables should have identical correlations with each group), the contribution of each input variable within the model must be statistically significant, and the distribution of the total sample must not be overly concentrated in one group. Table A6a depicts multiple comparison results between cluster means of each input variable to test for differentiation between clusters. In order to observe distinctions among groups with overlapping characteristics (e.g., comparing two groups in the same age cohort but differ in psychological or situational circumstances), we allowed for some input variables to not have statistically significant differences between a limited number of groups.

## Results

Greater risky behavior was indexed based on the average of items rated as having higher risk than benefit: ‘Risks are much greater than benefits’ or ‘Risks are somewhat greater than benefits’. Greater individual risk propensity was calculated as an individual’s regression between each item (except those labeled travel) and their rating of its riskiness relative to necessity or benefit, considering that under typical circumstances, these behaviors would be considered highly normative. Self-other gap in risk assessment was calculated as the difference between an individual’s rating of the potential harm to themselves and their rating of harm to others.

### Factor Analysis

We conducted an exploratory factor analysis to verify the structure of our measure COVID Risk Taking Inventory (CRI). The factor analysis used diagonally weighted least squares^48^, since reports of risk-taking behavior were ordinal (frequency of behavior), and an oblique rotation was applied to the factors due to the high correlation among some items^49^. A scree plot indicated that three or four factor models were equally well fit for capturing optimal variance. The four-factor model was deemed the most appropriate. The three-factor model loaded using public transit and carshare service items with other high-risk, low-frequency behaviors, such as meeting with a stranger in person for essential business, while the four-factor model included these items as a separate factor. Since we did not have participants report base rates for transit or use of car and/or carshare service, the travel factor was difficult to interpret. Thus, we excluded it from further analysis. The rank loadings are reported, along with sample variable descriptive statistics, in Table 1.

**Table 1.** Sample Descriptives of Study Variables

| **Input (Situational + Dispositional Clusters)** |  |  |  |  | **Output (Risk Behavior, Evaluation, Harm)** |  |  |  |  | **Factor Analysis** |  |
| --- | --- | --- | --- | --- | --- | --- | --- | --- | --- | --- | --- |
| <u>Descriptives</u> | <u>Mean</u> | <u>SD</u> | <u>Skew</u> | <u>Kurt.</u> | <u>Mean</u><br><u>Risk</u> | <u>Behavior</u> | <u>Eval</u> | <u>Self</u> | <u>Others</u> | <u>Label</u> | <u>Loading</u> |
| Living Space | 3.481 | 1.050 | -0.075 | -0.700 | Grocery | 3.680 | 2.790 | 2.380 | 2.780 | Essential | 0.621 |
| Perceived Scarcity | 4.274 | 1.699 | -0.334 | -0.910 | No Mask | 3.963 | 2.575 | 2.527 | 2.732 | Essential | 0.518 |
| Age | 37.124 | 11.318 | 0.958 | 0.048 | Friend at Distance | 3.616 | 2.465 | 2.367 | 2.786 | Low Risk | 0.476 |
| Extraversion | 28.853 | 7.663 | -0.436 | 0.402 | No Wash | 3.083 | 2.102 | 2.068 | 2.988 | Low Risk | 0.679 |
| Need for Cog. Closure | 60.834 | 10.604 | -0.463 | 0.223 | Stranger | 2.488 | 1.971 | 1.988 | 2.967 | High Risk | 0.680 |
| Behavioral Inhibition | 16.112 | 4.252 | 0.073 | 0.292 | Public Space | 2.602 | 2.133 | 2.083 | 2.927 | Low Risk | 0.614 |
| Empathy | 25.145 | 5.069 | 0.066 | -0.211 | Uber | 2.166 | 1.965 | 1.985 | 2.998 | Travel† | 0.860 |
| Conscientiousness | 40.276 | 8.068 | 0.075 | -1.034 | Exercise | 3.886 | 2.784 | 1.892 | 3.015 | Essential | 0.644 |
| Behavioral Activation | 27.861 | 7.099 | 0.306 | -0.279 | Transit | 2.270 | 1.971 | 2.697 | 2.558 | Travel | 0.821 |
| Openness | 43.033 | 7.423 | -0.954 | 1.352 | Event 5† | 2.23 | 1.917 | 1.925 | 3.039 | High Risk | 0.822 |
| Agreeableness | 38.417 | 7.143 | 0.316 | -0.387 |  |  |  |  |  |  |  |

† Travel variables were excluded from further analysis.

To label each factor, we used the item-level mean for activity risk assessment, which was determined by the overall sample ratings regarding risk-to-benefits (or necessity) for each activity. Activities labeled as *essential* included going to the grocery store, going outside without a mask, and exercising outside in public. *Low-risk* activities included returning home without washing hands, meeting a friend while maintaining social distance and visiting a public space. Finally, *high-risk* activities included attending a gathering with more than five guests not in your household, as well as interacting with a stranger for essential purposes. Another variable was used to estimate overall engagement in ‘Risky behavior’, which averaged all items from both low-risk and high-risk factors, and was deemed normally distributed using the Wilck-Shapiro test of normality and visual inspection. As expected, the low and high-risk factors were not normally distributed; non-parametric approaches were used to estimate the regression fit. Essential behavior and self-other ratings were also normally distributed.

### Shapley Regression

We applied a Shapley and OLS regression to examine the role of dispositional, situational and demographic variables in accounting for engagement in behavior deemed by the CDC ‘risky’ for spreading or being infected by the COVID-19^9^. First, all relevant ordinal variables (12-criterion model) were entered, accounting for 43% of the variance in risky behavior (see Table 2 for variable list). However, variables in this model were highly correlated – following the correlation output, we pruned variables for non-significance and multicollinearity violations (Conscientiousness was detected as a variance inflation factor greater than 10). The leaner, subsequent regression (7-criterion model) included Extraversion, Need for Closure, Empathy, Behavior Activation, Perceived Scarcity, Living Space size and Age. More frequent report of risky activity was associated with *higher* Extraversion, *greater* Need for Closure and Behavior Activation, as well as *more* perceived scarcity; in contrast, more frequent report of risky activity was associated with *lower* Empathy, *less* Living Space and *younger* Age. Since this set of criterion variables were relevant in predicting variability in behavior, we proceeded to check these for normality violations in order to submit those in the 7-criterion model for submission to a stepwise regression model.

**Table 2.**
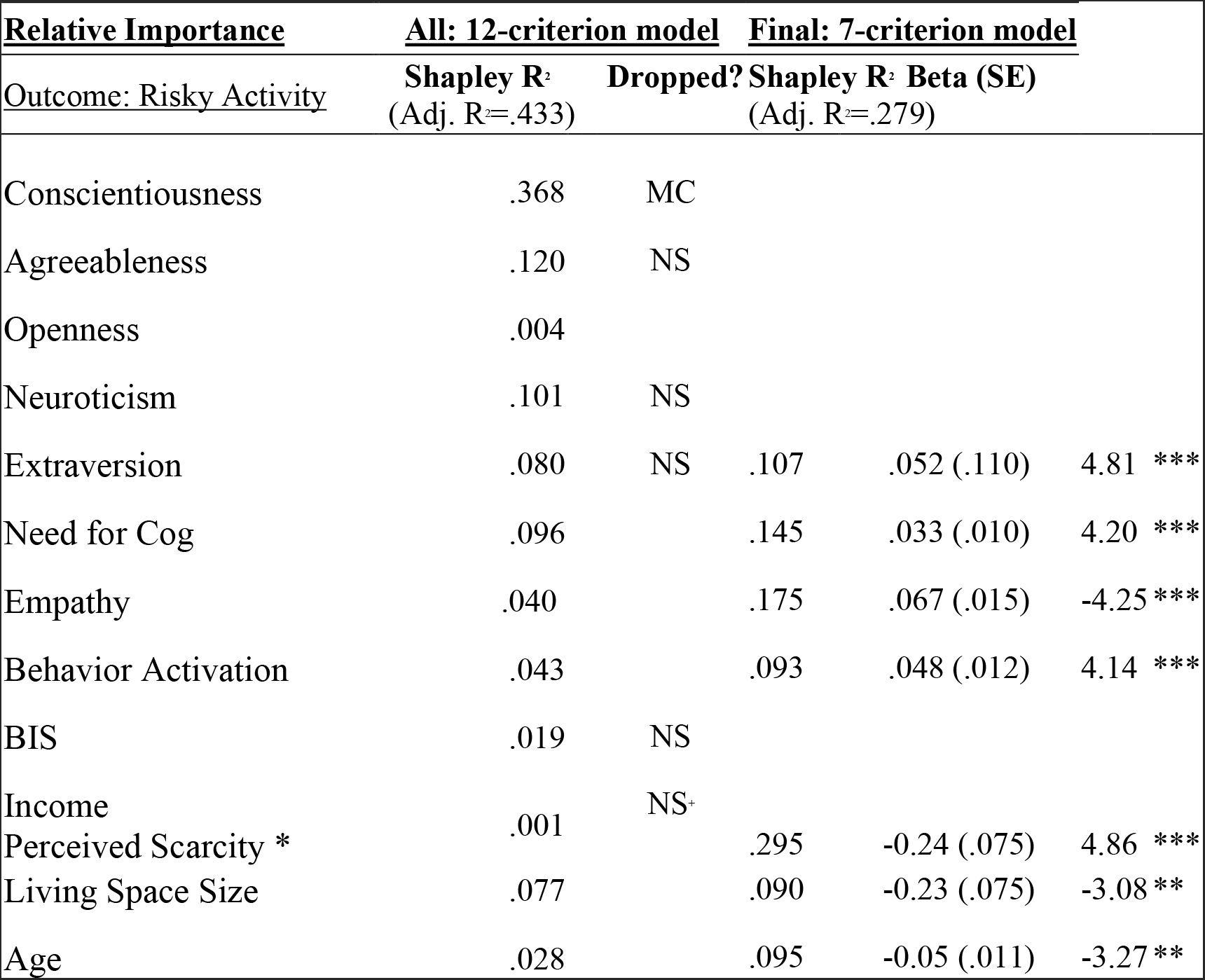
Shapley Regression: Examining Predictors of Risky Activity.

Sample-wide descriptive statistics and correlations are provided in Table 3. Dispositional and situational factors were examined visually and using the Wilck-Shapiro test for normality; all were normally distributed. Since situational variables related to employment (e.g., whether participants select the statement “I am classified as an essential worker”) were not asked for the full sample (they were follow-up questions to individuals who identified themselves as employed or unemployed, a binary), we did not consider them as input variables for the cluster model and tested them separately from other situational variables. In addition, we note that 52 out of 482 individuals reported that they are not currently employed. Of the participants who were employed (430), 39% were able to continue working remotely and 17% identified themselves as ‘essential’ workers. Participants reported spending 36.47% of their time outside their home for employment.

**Table 3.** Cumulative Model: Risky behavior by Stepwise Demographic, Situational and Dispositional Variables.

| Model |  | Unstandardized | Standard Error | Standardized | t | p |
| --- | --- | --- | --- | --- | --- | --- |
| 1 | (Intercept) | 0.347 | 0.653 |  | 0.532 | 0.595 |
|  | Employ | -0.038 | 0.014 | -0.121 | -2.740 | 0.006 |
|  | Income | -0.117 | 0.055 | -0.094 | -2.135 | 0.033 |
|  | Education | 0.199 | 0.056 | 0.159 | 3.534 | < .001 |
|  | Political Orientation | 0.272 | 0.040 | 0.288 | 6.787 | < .001 |
|  | Gender | 0.376 | 0.123 | 0.128 | 3.053 | 0.002 |
| 2 | (Intercept) | -1.056 | 0.746 |  | -1.415 | 0.158 |
|  | Employ | -0.037 | 0.014 | -0.118 | -2.700 | 0.007 |
|  | Income | -0.119 | 0.054 | -0.096 | -2.199 | 0.028 |
|  | Education | 0.195 | 0.056 | 0.156 | 3.511 | < .001 |
|  | Political Orientation | 0.257 | 0.040 | 0.272 | 6.477 | < .001 |
|  | Gender | 0.399 | 0.122 | 0.136 | 3.281 | 0.001 |
|  | Need for Closure | 0.022 | 0.006 | 0.155 | 3.725 | < .001 |
| 3 | (Intercept) | 0.035 | 0.895 |  | 0.039 | 0.969 |
|  | Employ | -0.038 | 0.013 | -0.122 | -2.808 | 0.005 |
|  | Income | -0.118 | 0.054 | -0.095 | -2.186 | 0.029 |
|  | Education | 0.177 | 0.056 | 0.141 | 3.153 | 0.002 |
|  | Political Orientation | 0.241 | 0.040 | 0.255 | 5.978 | < .001 |
|  | Gender | 0.358 | 0.123 | 0.122 | 2.917 | 0.004 |
|  | Need for Closure | 0.022 | 0.006 | 0.151 | 3.639 | < .001 |
|  | Empathy | -0.029 | 0.013 | -0.094 | -2.190 | 0.029 |

In a separate ANOVA, there were no significant differences overall in risky behavior by race, or within dummy-coded racial identity categories. See appendix A2 for racial composition of sample. Our model does not allow enough df to test race properly with dummy-codes, so we excluded it for now.

### Multiple Regressions

We ran a series of multiple regressions, with the goal of examining whether the dispositional and situational variables contributed to significant variance to a range of COVID-19 risk-taking behavior.

Dispositional factors alone accounted for 10.1% of the variance in risk taking, F (4, 477) = 14.473, p < 0.001. Situational factors alone accounted for 14.7% of the variance in risk taking, F (3, 478) = 8.939, p < 0.001. Demographic factors alone accounted for 17.6% of the variance in risk taking, F (10, 471) = 12.518, p < 0.001. Controlling for situational factors, dispositional factors accounted for an additional 7.2% of the variance in risk taking, F (7, 474) = 9.725, p < 0.001.

Engagement in high-risk activities was associated with all dispositional factors, F (7, 474) = 24.916, p < 0.001, R^2^ = .269, controlling for all situational factors, F (3, 478) = 31.692, p < 0.001. All dispositional factors were significant, accounting for a further 10.3% of the variance in high-risk activity engagement. It would appear younger individuals and those reporting greater perceived scarcity engaged in more frequent high-risk activities, particularly those who were more extraverted, higher in need for cognitive closure and sensation-seeking, and lower in empathy.

Engagement in low-risk activities was associated with dispositional factors, F (7, 474) = 14.371, p < 0.001, R^2^ = .163, controlling for situational factors, F (7, 474) = 14.497, p < 0.001. Specifically, more frequent low-risk activity was significantly associated with greater extraversion, need for cognitive closure and less empathy, accounting for a further 9.2% of the variance. Engagement in essential activities was situational factors, F (7, 474) = 3.538, p = 0.015, R^2^ = .028 and not further with dispositional factors, F (3, 478) = 2.989, p = 0.059. Specifically, more frequent essential activity was significantly associated with more living space and higher perceived scarcity. Regression results for low-risk and essential activities are shown in Table 3A and 4A in the appendix.

**Table 4.** Summary of Cluster Groups based on Input Variables and Demographic Differences.

| **Cluster Groups** | ***Psychological & Situational Summary based on input variables*** | ***Demographic and Quarantine Compliance Summary (based on select output variables, <u>see Appendix for corresponding data</u>).***<br>***SA = Sample Average*** |
| --- | --- | --- |
| ***Cluster 1 (n=53)*** | Middle in terms of introversion and highest in age. Also moderate in sensation seeking and perceived scarcity. Lower on empathy and not likely to have large living space. | Skews low income (42% vs 35% SA) and majority live in suburbs (55%). Also leans towards Conservative (57%). Shows lower levels of trust toward organizations like CDC compared to other clusters (70% vs 81% SA), but still remains cautious against COVID. More likely to get COVID information from cable news relative to other clusters. Least likely to say that going to a public space or using public transit is high risk. Most likely to say that employment status did not change since COVID (67% vs 57% SA). Skews mainly White (87%). |
| ***Cluster 2 (n=58)*** | Highly introverted and likely to live in a large living space. Still fairly young and very low in sensation seeking. Lower-middle end in empathy. Has low levels of perceived scarcity during the pandemic. | Younger, largely skews Millennial and Gen Z. Least likely to be employed full-time (55% vs 75% SA). Mostly Liberal (67%) and majority live in urban or suburban areas (90%). Takes quarantine seriously and is supportive of CDC. Complies with quarantine guidelines but does occasionally go outside for leisure despite being the least likely to agree that going outside for non-essential needs is ok (29% vs 48% SA). |
| **Cluster 3**<br>**(n=93)** | Not too introverted and very young. Scores low in both sensation seeking and empathy. Not likely to live in a large living space and has scores moderately high in perceived scarcity. | Most rural relative to other Clusters (33% vs 22% SA). Majority middle income but also skews lower (94% earn less than \$99,999 a year). Leans Conservative (64%) and more likely to have children relative to other clusters (60% vs 49% SA). Recognizes risks of COVID but generally does not practice strict quarantine compliance. Has been impacted by COVID financially and has trouble finding resources. Still takes Uber and public transit, but might be related to higher likelihood of being an essential worker (25% vs 16% SA). Majority White (58%) but also skews Black American (29% vs 17% SA). |
| **Cluster 4**<br>**(n=109)** | Low introversion and very high in sensation seeking. Middle age range with moderate levels of perspective-taking empathy. Some live in a larger living space, but many others do not. Reports the highest in perceived scarcity. | Mostly middle income (62%) but skews lower income as well (31%). More likely to be Conservative (63%). Majority live in urban areas (41%) but many also live in rural and suburban regions. Middle age and most likely to have kids (62% vs 49% SA). Respects authority figures like CDC and takes them seriously but also very lax on complying with quarantine. Believes that a lot of restrictions are excessive. Most likely to be an essential worker (28% vs 16% SA) and also be impacted financially by COVID. Majority White (61%) but also skews Black American (31% vs 17% SA). |
| **Cluster 5**<br>**(n=41)** | Lowest in introversion and the youngest cluster. High levels of sensation seeking and fairly empathetic. Not too likely to live in a larger living space but also reports fairly low in perceived scarcity. | Most likely to be working full-time (93% vs 75% SA). Majority are Liberal (63%) and Millennial / Gen Z. Over 90% live in either urban or suburban areas. Respects quarantine compliance and mainly goes outside for essential needs. Reports the lowest amount of hours outside for leisure reasons (2.45 vs 5.28 SA). Sees risk in public spaces and generally avoids them. |
| **Cluster 6**<br>**(n=75)** | High in both introversion and age. Very likely to live in a larger living space but reports moderately high in perceived scarcity. Fairly low in sensation seeking with moderate levels of empathy. | Majority live in Suburbs (55%) but also skews rural (27%). More likely to have low income relative to other clusters (47% vs 35% SA). Political orientation is fairly spread out. Very wary of COVID threat and perceives many activities as high risk. Generally avoids going outdoors. Has been impacted financially by COVID but still complies to quarantine. |
| **Cluster 7**<br>**(n=53)** | Moderate in introversion but very high in age, sensation seeking, and perspective-taking empathy. Most likely to live in a large living space and scores very low in perceived scarcity. | Most likely to have 100k+ income compared to other clusters (28% vs 15% SA). Liberal leaning (23% Very Liberal vs 15% SA; however only 53% are Liberal overall) and more suburb/rural centered (77%). Compliant to quarantine and perceives many non-essential activities as risky. Main reason to go outside is for employment. Respects the CDC and WHO the most out of the other clusters. Least likely to say that COVID pandemic has impacted them financially (25% vs 53% SA). Rarely uses public transit, but might be because they have access to other modes of transportation. Least racially diverse: 89% White. |

Next, we ran a stepwise regression on the dispositional (Extraversion, Need for Cognitive Closure, Empathy, Behavior Activation) and situational (Age, Living Space Access and Perceived Scarcity) variables identified by the lean Shapley regression, as well as the six demographic variables (Income, Education, Employment, Political Orientation and Gender – Race was not included^1^) significantly associated with risky behavior, as indicated by preliminary correlations (see Appendix 5). In the stepwise model, variables were nominated as accounting for Risky behavior in the following sequence: Political Orientation, Education, Need for Cognitive Closure, Gender, Empathy and Income. Other variables, including Employment, were not nominated.

Subsequently, to examine if significance remained after controlling for demographic characteristics, we used a hierarchical linear model (Table 3). Our analyses indicated that block of situational (3%) and dispositional (7%) variables contributed unique significance to variance in risky behavior, relative to the demographic (10%) variables. Specifically, dispositional variables of Need for Cognitive Closure (5%) and Empathy (2%) individually held their significance, controlling for demographic variables, although no other situational or dispositional variables remained individually significant.

### Cluster Analysis

Groups for the analysis were created using *k*-means clustering using variables related to psychological and situational circumstances. The final input variables used in the model reflected those ranked by the learn Shapley regression in relation to risk propensity: introversion scores measured by the BFI, sensation seeking scores measured by BIS/BAS, perspective-taking empathy scores from the IRI scale, age, living space access (whether or not the participant lives in a residence with more than 2 common spaces), and perceived scarcity (how much participants agree with the statement “it has been difficult for me to get needed resources (food, toilet paper) due to the Coronavirus” (1 Strong disagree to 7 Strongly agree). To ensure that there was adequate differentiation between clusters, only models that showed a majority of statistically significant comparisons were considered. Additionally, an ANOVA was conducted on input variables to assess significant contribution of each variable. Complete results from the cluster analyses are presented in Table A6b of the appendix. Table 4 shows high level summaries for each cluster, based on both input and selected outcome variables. See Table A1 in the appendix for a more detailed look at demographic and behavioral variables related to each Cluster. Table 5 shows the percentage of each cluster that at least somewhat agrees with statements that reflect attitudes towards quarantine restrictions.

**Figure 1.**
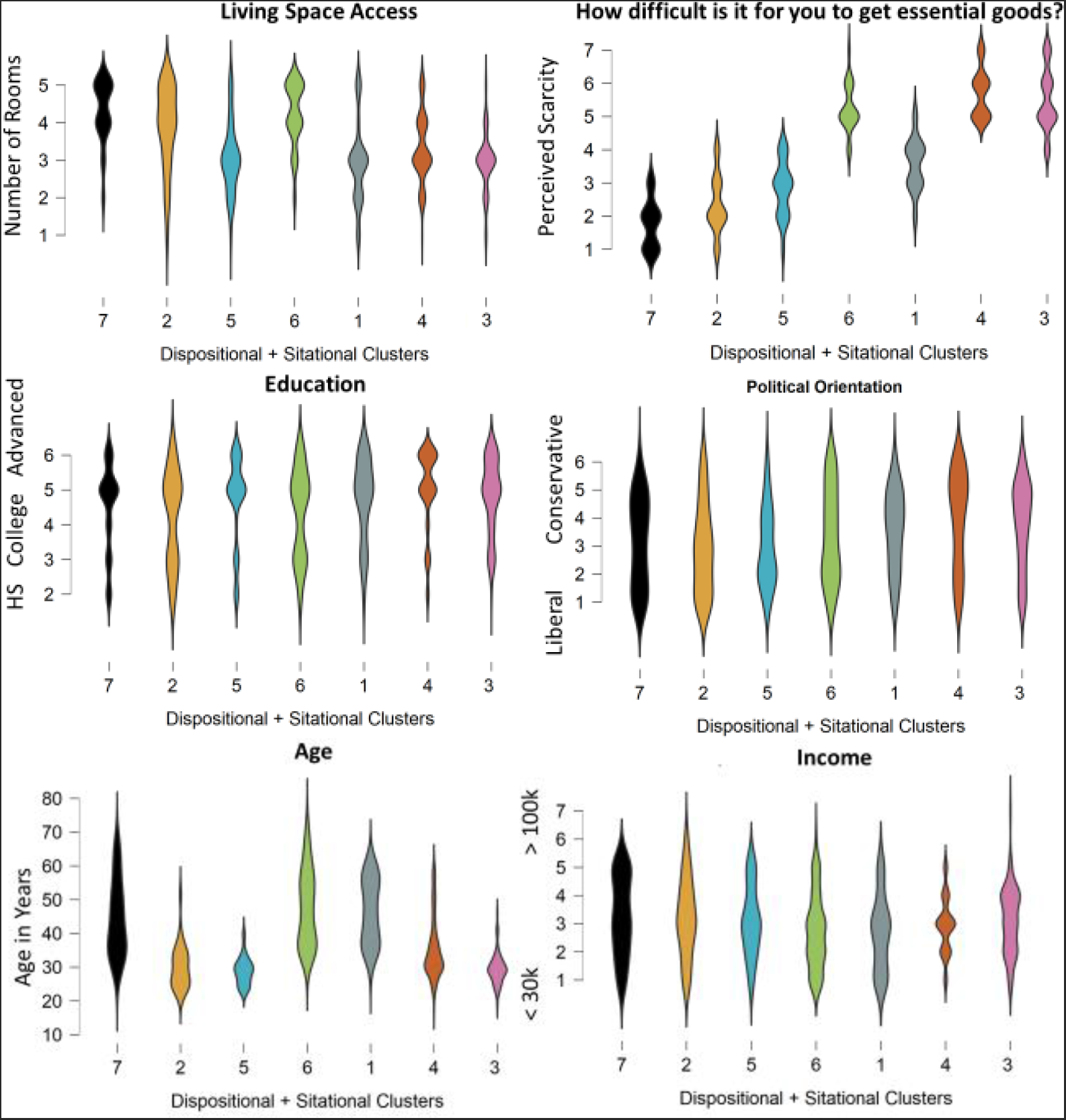
Examining Cluster Differences in Situational and Demographic Features. Violin plots display the distribution and range of values in each of seven clusters, which were parsed by dispositional variables and situational variables (Living Space Access and Perceived Scarcity) as well as Age. As shown, clusters differed on other demographic features, specifically education, income and education.

**Table 5.**
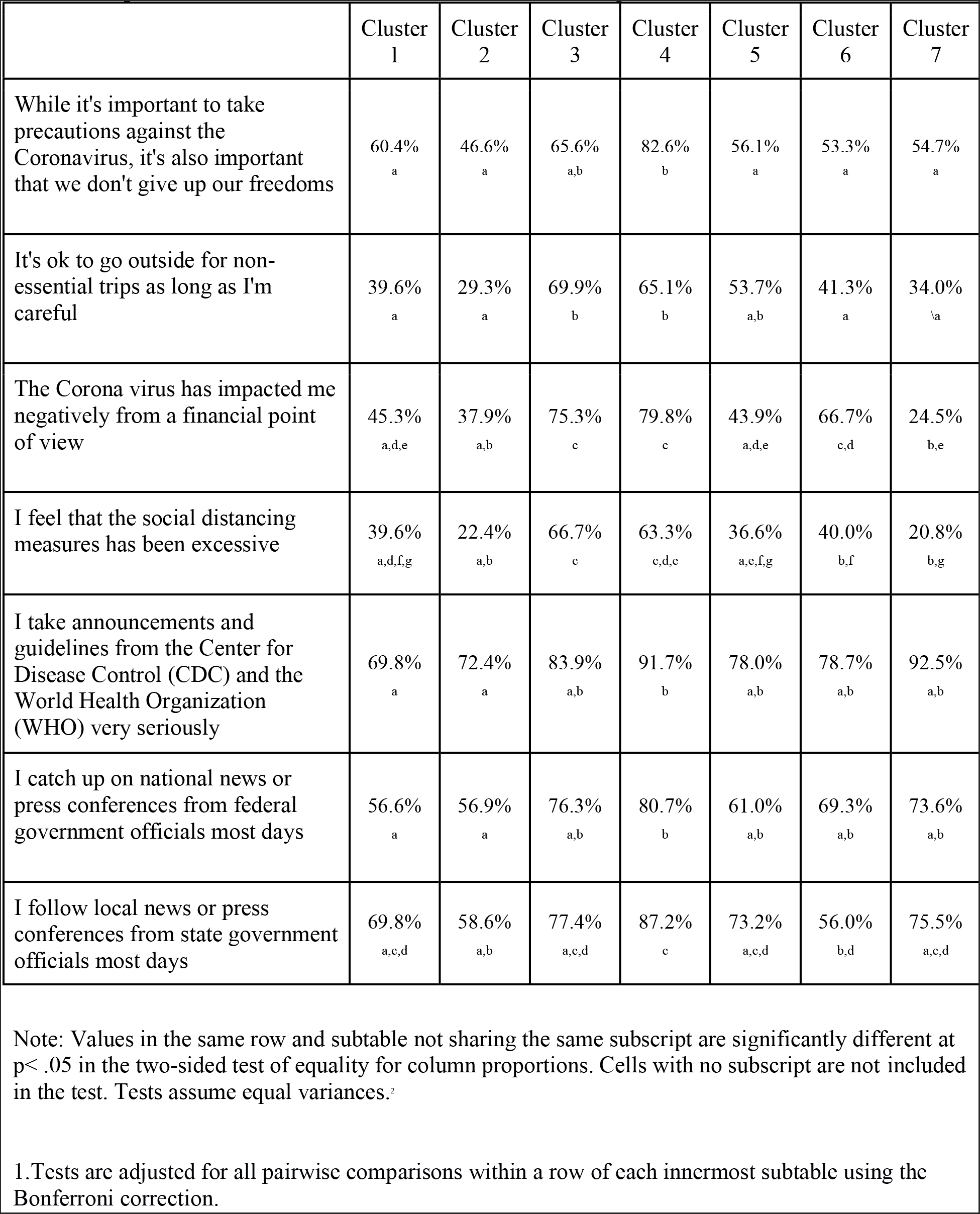
Proportion of each Cluster that at least “Somewhat Agrees” to each statement.

### Risky behavior and activity by Cluster

We identified the risk taking propensity of clusters by first providing each subject with a regression score for their risk taking, given their subject-specific cost-to-benefit evaluation. The clusters were ranked 1–7 by the proportion of individuals who engaged in activities they rated as ‘risks outweigh the benefits’ – this is the order in which they are subsequently presented. A triadic split was applied to discriminate which clusters had the lowest (clusters 7 and 2) and highest risk taking propensity (clusters 4 and 3), and labeled them as ‘risk averse’ and ‘risk inclined’ respectively. Moderate risk-takers were labeled as ‘compliant’ (clusters 5, 6 and 1).

Using ANOVA, we examined cluster differences in frequency of activity engagement across all CRI items (overall activity) and three factors of risk activities, determined by the factor analysis. Overall activity differed by cluster, F (6) = 3.062, SS = 40.387, p = 0.006, ω² = 0.025, such that individuals belonging to high-risk clusters 3 and 4 reported engaging more frequently in all activities, particularly evident relative to cluster 7. High risk activity differed by cluster, F (6) = 14.974, SS = 48.657, p < 0.001, ω² = 0.148, such that individuals belonging to cluster 3 reported engaging more frequently in high-risk activities relative to all other individuals (the significance was marginal when compared to individuals from high-risk clusters, 3 and 4). Individuals belonging to risk-averse cluster 7 engaged in significantly less high-risk activity relative to the individuals in clusters identified as compliant and high-risk. Low-risk activity differed by cluster, F (6) = 7.365, SS = 23.364, p < 0.001, ω² = 0.073, such that individuals belonging to clusters 3 and 4 reported engaging more frequently in low-risk activities relative to individuals in all other clusters. Individuals belonging to risk-averse cluster 2 engaged in significantly less activity relative to the individuals in clusters identified as compliant and high-risk. Essential activities did not differ by cluster, F (6) = 1.286, SS = 18.761, p = 0.262, ω^2^ = .002.

### Employment Related Situational Variables and Cluster Group Comparisons

Using bivariate correlations, we examined associations between situational variables related to participant’s employment circumstances and frequency of high-risk activities. Employment-related variables include: employment status, the percentage of total working hours spent on a job site or surrounded by other people, classification as an essential worker, and whether or not the participant is able to work remotely. The results below show that being currently employed, percentage of total working hours on a job site, and being classified as an essential worker are positively correlated with both high-risk activities. These effects are also statistically significant at p < .01. Being able to work remotely shows negative correlations with both high-risk activities, however only the correlation with meeting in-person with a stranger is significant.

**Table 6.**
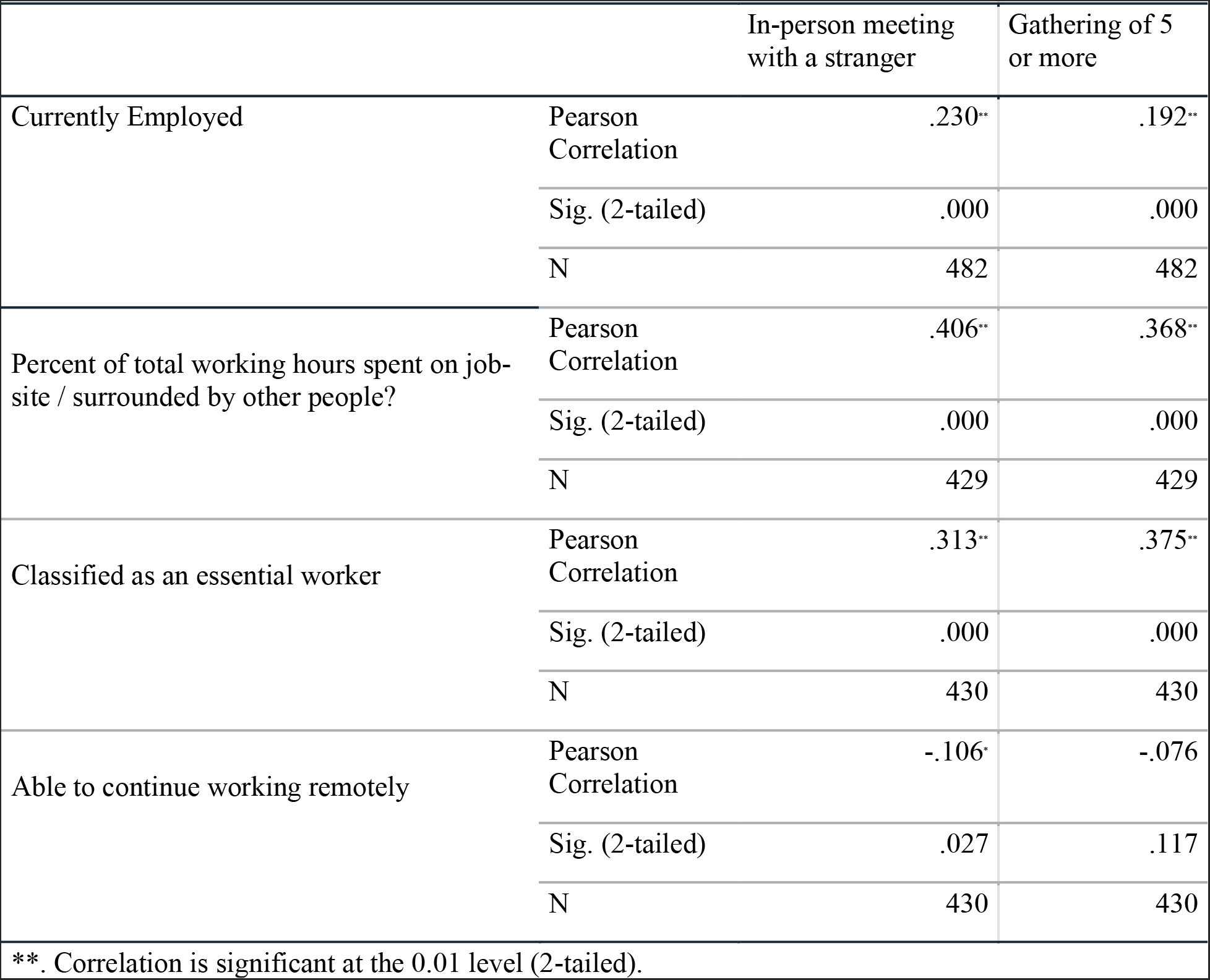
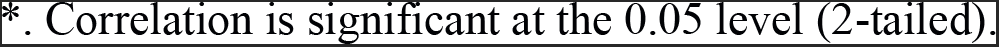

|  |  | In-person meeting<br>with a stranger | Gathering of 5<br>or more |
| --- | --- | --- | --- |
| Currently Employed | Pearson<br>Correlation | .230** | .192** |
|  | Sig. (2-tailed) | .000 | .000 |
|  | N | 482 | 482 |
| Percent of total working hours spent on job-site / surrounded by other people? | Pearson<br>Correlation | .406** | .368** |
|  | Sig. (2-tailed) | .000 | .000 |
|  | N | 429 | 429 |
| Classified as an essential worker | Pearson<br>Correlation | .313** | .375** |
|  | Sig. (2-tailed) | .000 | .000 |
|  | N | 430 | 430 |
| Able to continue working remotely | Pearson<br>Correlation | -.106* | -.076 |
|  | Sig. (2-tailed) | .027 | .117 |
|  | N | 430 | 430 |

\*\*. Correlation is significant at the 0.01 level (2-tailed).
\*, Correlation is significant at the 0.05 level (2-tailed).

Once we identified that employment-related situational variables were associated with frequency of high-risk activities, we ran another set of bivariate correlations between cluster groups and employment variables. In this analysis, we intended to observe how the Risk-inclined groups (clusters 3 & 4) and Risk-Averse groups (clusters 2 & 7) differ in relation to situational variables that are correlated with increased engagement of high-risk activities. As depicted in Table 7, both Risk-inclined and Risk-Averse cluster groups show statistically significant correlations with all employment-related variables except for ability to work remotely. However, Risk-inclined groups show positive correlations with employment variables, while Risk-Averse groups are negatively correlated. This suggests that Risk-inclined groups are more likely to be involved in work-related situations that are positively correlated with increased frequency of high-risk activities.

**Table 7.**

|  |  | Currently employed | Percent of total working hours spent on job site / surrounded by other people? | Classified as an essential worker | Able to continue working remotely |
| --- | --- | --- | --- | --- | --- |
| High Risk Clusters (3 & 4) | Pearson Correlation | .146** | .277** | .174** | -.023 |
|  | Sig. (2-tailed) | .001 | .000 | .000 | .641 |
|  | N | 482 | 429 | 430 | 430 |
| Cluster 3 | Pearson Correlation | .102* | .148** | .080 | -.038 |
|  | Sig. (2-tailed) | .025 | .002 | .096 | .427 |
|  | N | 482 | 429 | 430 | 430 |
| Cluster 4 | Pearson Correlation | .076 | .182** | .127** | .010 |
|  | Sig. (2-tailed) | .095 | .000 | .008 | .832 |
|  | N | 482 | 429 | 430 | 430 |
| Risk-Averse Clusters (2 & 7) | Pearson Correlation | -.143** | -.127** | -.114* | -.009 |
|  | Sig. (2-tailed) | .002 | .009 | .018 | .855 |
|  | N | 482 | 429 | 430 | 430 |
| Cluster 2 | Pearson Correlation | -.118** | -.061 | -.049 | .025 |
|  | Sig. (2-tailed) | .009 | .208 | .306 | .600 |
|  | N | 482 | 429 | 430 | 430 |
| Cluster 7 | Pearson Correlation | -.070 | -.108* | -.102* | -.038 |
|  | Sig. (2-tailed) | .124 | .025 | .034 | .435 |
|  | N | 482 | 429 | 430 | 430 |

\*\*. Correlation is significant at the 0.01 level (2-tailed).
\*. Correlation is significant at the 0.05 level (2-tailed).

### Exploratory: Self-Other Risk Assessment by Cluster

We examined whether the clusters differed according to whether they assessed riskiness as greater for themselves relative to others. This score was obtained by subtracting perceived risk to others from perceived risk to self, such that more negative scores indicate greater risk estimated to others. The initial one-way ANOVA was not significant, (6) = 1.480, SS = 9.035, p = 0.183, ω² = 0.006. In the absence of linear effects and in light of the visualization (figure 4), we probed further using non- parametric paired contrasts. These yielded significant quadratic effects: differences relied on the ordinal ranking of cluster by risk propensity, suggesting that individuals belonging to the moderate ‘compliant’ were more likely to report significantly more negative self-other risk assessment relative to the sample average. More negative values indicated more concern for harm to others than to themselves. In contrast, risk-averse and risk-inclined individuals were more likely to report smaller gaps between concern for harm to themselves and others.

**Figure 3.**
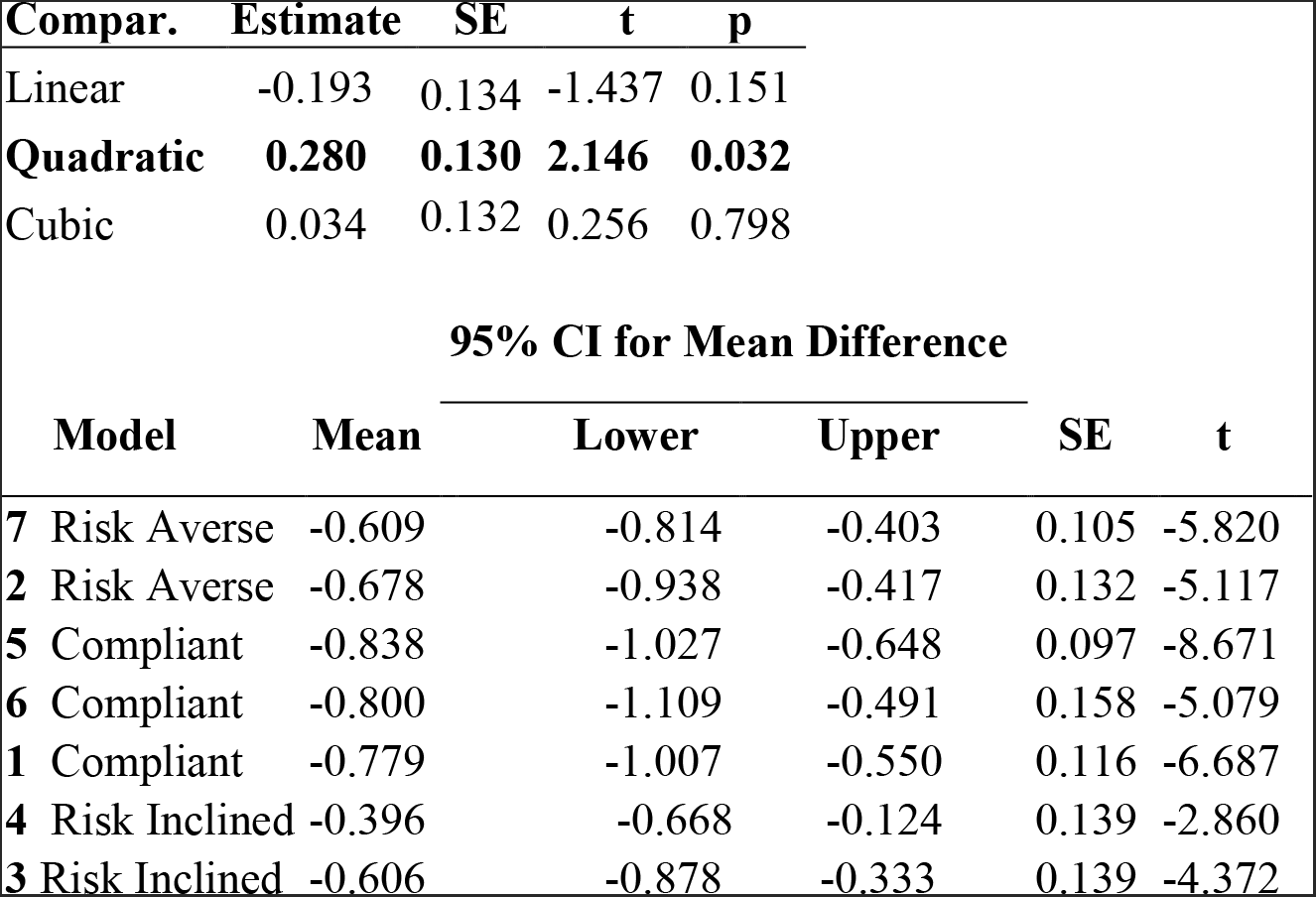
Change high-risk to risk-inclined (to prevent confusion).

## Discussion

Our initial goal was to examine whether the dispositional variables contributed to significant variance for a range of COVID risk-taking behaviors, controlling for variance accounted for by situational variables. More frequent report of risky activity was associated with *higher* extraversion, need for cognitive closure, behavior activation, and perceived scarcity, but *less* empathy and living space access, as well as *younger* age. Our regression analyses offered a sample-wide view of these trends, associating situational and dispositional attributes with propensity for risk taking and risk assessment. The cluster analysis yielded profiles of ‘types’ of individuals, hinting toward candidate motives and explanations for overall non-compliance. Combined with testing differences in risk taking by cluster identity, the profile approach allows consideration of multi-faceted attributes, which influence an individual’s receptiveness to adhere to public health measures, responsiveness to health promotion and outbreak communication guidance, and ultimately impact the effectiveness of COVID-19-related interventions.

To gain insight into the personal attributes and contexts that contribute to CDC guideline adherence, we probed the significance of these apparent differences in risk taking and general activity by dispositional-situational clusters. In addition to an effect for overall activity and risky activity, we found significant differences by cluster in low-risk and high-risk activities, which are subscales of the CRI identified using sample-specific ratings of the risk-to-benefit (or necessity). There were no significant differences in essential activity by cluster analysis, which further corroborates the selectivity of cluster input variables. We suggest that extending beyond simple demographics, contextual and psychological tendencies allows for the discrimination of risky behavior from ordinary or necessary activity. For our sample, 42% of participants were identified based on their situational-dispositional cluster as risk-inclined compliant (and in fact engaged in more frequent risk taking, especially for high-risk activities like attending gatherings and meeting with a stranger). While we do not claim that this proportion is reflective of the actual proportion of US citizens engaging in high-risk activities due to sample restraints, it appears that while the new ‘normative’ behavior is to comply with public health guidance there is a sizeable proportion of individuals who may not be able or willing to adhere to certain recommendations.

Finally, an exploratory analysis indicated that self-other risk evaluation might have a non-linear association with cluster risk engagement, which was ranked by risk tendency. Across all items, participants consistently viewed the perceived risk to other as greater to themselves, except for the use of public transit. We found a quadratic effect, such that individuals identified as normative compliant (the majority of participants) rather than risk-averse compliant (clusters 7 and 2, the lowest in risk behavior) or risk-inclined compliant (clusters 3 and 4, the highest in risk behavior) had the smallest gap in their assessment of risk to self-versus risk to others. It is conjectured that while most individuals are influenced by public health messaging aimed to invoke consistency in community response, perhaps at the expense of individual choice, the most risk-averse individuals may be motivated by protecting their own health. Similarly, individuals who might otherwise be risk-inclined compliant or even actively non-compliant with CDC guidelines might be more responsive to public health messaging if they feel an activity might have a personal cost to them, beyond the utility of the community benefit of adherence.

### Profiles of compliance from dispositional and situational factors

From conducting k-means clustering we produced a model with 7 cluster groups based on psychological and situational variables. In a social distancing context, cluster 1 remains compliant yet cautious towards authority, reporting less trust towards organizations like the CDC. Cluster 2 is a younger group that reports high adherence to social distancing guidelines, both in how frequently they engage in outside activities and attitudes towards the need to quarantine (however, they also report going outside occasionally for leisure). Cluster 3 is a younger group who appear to be the least empathetic and most likely to live in a rural area, relative to the other clusters. Members of cluster 3 show engagement in high-risk activities, and are more likely to agree with statements such as “I feel that the social distancing measures have been excessive” and “It’s ok to go outside for non-essential trips as long as I’m careful.” Cluster 4 shows similar non-compliance attitudes and risk-inclined behaviors, despite being older and more sensation-seeking than cluster 3. However, it is noteworthy that both clusters 3 and 4 are more likely to be classified as essential workers, and report spending higher percentages of their working hours outside or surrounded by other people. They also report the highest agreement with the statement “The Coronavirus has impacted me negatively from a financial point of view” compared to all other clusters. This difference is statistically significant as well. Cluster 5 is the youngest cluster and also the most likely to be working full-time (up to 93%). This cluster demonstrates high compliance with quarantine measures and reports going outside for leisure the least among the other groups. Cluster 6 is an older group that perceives many outdoor activities as risky and mostly avoids the outdoors. Cluster 7 is another older by public health messaging aimed to invoke consistency in community response, perhaps at the expense group, but has the highest average income relative to the other clusters, with higher levels of social distancing compliance in both attitude and behavior. Cluster 7 is also the least likely to report that the Coronavirus has impacted them negatively from a financial point of view (24.5% vs 75.3% and 79.8% reported by clusters 3 and 4 respectively).

## Limitations

While samples obtained from MTurk are widely used in the social sciences^35^, it is important to note that a sample obtained in this way may not be nationally representative of the American response to COVID-19. In addition, while our sample size was sufficient to detect even subtle effects, all measures reported here were self-report, and thus, biases related to self-report could be a confounding factor limiting the specificity of any insights into effects on public health compliance. In follow-up work, our CRI measures could be complimented by smartphone kinematic and geolocation data, as well as indicators of mobile and digital device use more broadly – these measures have been used to indicate behavioral compliance and frequency of information consumption related to public health guidance^34^. Further, given the remote nature of their employment, MTurk respondents are more likely to have more computer experience and multiple forms of employment than the typical American population. Thus, our sample is likely underestimating the impact of dispositional and situational factors on psychological state and risk taking during COVID-19.

It is possible that the unusual circumstances of the COVID pandemic and social distance measures might impact responses as well. In fact, the heightened risk perception rating of routine activities is an indicator of the unusual, time-sensitive nature of our results. The origins of variability in COVID-19 epoch risk patterns observed are also difficult to disentangle from typical variation in risk propensities across our current sample, since we had no assessment of base rates of behavior or prior risk ratings. Further, though we cannot extricate our sample from those exposed to mortality threats related to COVID-19, our study might be considered an example of how mortality salience differentially influences risk engagement and attitudes, such that differences in disposition have been associated with self-protective and self-destructive tendencies^50^. Future studies designed to experimentally manipulate (rather than study individual differences in) death salience will be useful in identifying whether the variability observed in risky behavior in the present study is a distal or proximal mechanism related to theories of mortality-motivated changes in risk behavior.

## Implications

The results from this study show that psychological and situational factors impact risk-taking behavior, which could result in non-compliance with social distancing guidelines. Our cluster model shows that groups exhibited a wide array of behaviors and attitudes towards the COVID-19 pandemic, indicating that underlying psychological and situational factors could drive some variability in behavioral compliance with public health guidance. Our analysis of risk-taking behavior also identified two ‘risk-inclined’ groups (clusters 3 and 4) that exhibited a higher propensity for engaging in currently high-risk activities. Members of both these groups were more likely to be conservative-leaning and reported high perceived scarcity of goods, suggesting a potential political agenda for their risk engagement and attitudes; however, these were distinct populations, who differed significantly in age, sensation-seeking, and region density. Upon further investigation, we found that these risk-inclined clusters were more likely to report circumstances relevant to their exposure (employment, perceived scarcity, and limited living space), along with their higher engagement in high-risk activities. It is possible that engagement in high-risk activities was partially driven by the circumstances that these individuals have encountered, and may not be solely due to demographic factors and psychological dispositions. This interpretation is also supported when examining risk-averse groups such as clusters 2 and 7, who avoided high-risk activities but were also less likely to be affected by these situational factors.

In general, since situational circumstances may make individuals more vulnerable to contracting COVID-19, and in many cases are outside the direct control of these individuals, these factors may be intractable issues that significantly impair the ability for certain population groups to adhere to stay at home orders and other outbreak measures. For example, if clusters 3 and 4 are the most likely to be essential workers and have to work on a job-site, then it is not surprising that they also engage in more high-risk activities compared to clusters 2 and 7, who are less likely to work around people. Clusters 2 and 7 are also the least likely to report that the COVID-19 pandemic has impacted them financially while clusters 3 and 4 are the most likely, which suggests that the groups who engage more frequently in non-compliant behavior are also the ones who are most vulnerable to the pandemic. Further, in evaluating demographic comparisons in infection and social distance compliance by racial compositions, ‘exposure’ and situational factors should be considered. For example, Black Americans who are more susceptible to contracting COVID-19 also have higher representation in clusters 3 and 4. As defined by Farmer et al. (2006)^51^ in their discussion of how public health practitioners can address social determinants of disease, ‘structural violence’ occurs when political, economic, and cultural structures are organized in ways that put individuals and populations in harm’s way. In terms of structural violence, it is likely that economic and political systems that existed before the COVID pandemic is still be impacting disparities in health and public health guidance adherence within this new context.

Based on these results, we recommend that circumstantial differences be considered when communicating social distancing guidelines, and also when developing policy approaches to ensure equitable and targeted outbreak control and response. Put simply, current quarantine, stay at home, and social distancing measures may not adequately take into account the complex cooccurrence of dispositional and situational factors that drive both compliance and non-compliance among the public. The combination of these characteristics may be independent of demographic factors often identified as barriers or enablers for health behavior and should be taken into account in public outreach. Our dispositional-situational clusters also provide initial insights into the combination of characteristics that may influence receptivity to health messages that can then be tailored to specific individuals based on initial survey data such as what was conducted in this study.

To improve compliance with public health guidance and develop messaging appropriate for different people under varying circumstances, we advise further consideration of the dispositional and situational factors influencing risk taking during COVID-19. In the same way that abstinence only education programs are correlated with higher rates of teen pregnancy^52^ and “Just Say No” drug prevention programs could lead to even higher drug usage^53^, it is possible that recommending guidelines that urge people to stay home could induce feelings of confinement, rebellion or mortality salience, leading to greater risk-taking behavior. Further, multivariate people-centered approaches to analyses, which consider both dispositional and situational variability, are advised for examining the efficacy of public health interventions. Messaging can be tailored for ‘types’ of audience, to further assess the utility of language used for persuading the public and in laws enforcing public health guidelines. There is a need for these types of assessments to be scaled for a larger number of respondents, to include a more generalized cohort representative of the U.S. population. In light of the recent reconceptualization of risk as a matter of routine, our findings contribute to a more nuanced understanding of how individual differences in attitudes and circumstances influence risk perception. Moreover, these outcomes show that the wider deployment of profile-centric approaches to assess public health compliance and messaging efficacy may be useful in critical efforts to “flatten” the COVID-19 epidemiological curve.

## Data Availability

The data has been submitted to the reviewing journal; it will be included as a supplement hosted by the journal if accepted for publication.

## Acknowledgements

We thank the participants for their willingness to share their time, disclose their behavior and reflect on their responses to COVID-19 during these uncertain times. We also thank Bryana Walker who’s conversations with one of the first authors helped shape the conceptualization of the study during its early stages.

## Appendix

**Table A1.** Demographic Breakdown of Full Sample (column percentages)

|  |  | Full Sample (n=482) |
| --- | --- | --- |
| *Gender* | Female | 39.8% |
|  | Male | 59.1% |
|  | Other | 0.6% |
|  | Prefer not say | 0.4% |
| *Age* | Less than 29 | 21.0% |
|  | 29-33 | 27.4% |
|  | 34-43 | 26.1% |
|  | 44+ | 25.5% |
| *Race* | White (Caucasian) | 71.2% |
|  | Black | 19.7% |
|  | Asian or Pacific Islander | 6.0% |
|  | American Indian or Alaskan Native | 1.2% |
|  | Hispanic | 7.1% |
|  | Non-hispanic | 0.4% |
|  | Other, specify: | 0.8% |
| *Income* | Under \$30,000 to Less than \$49,999 | 35.1% |
| | Between \$50,000 and \$99,999 | 51.5% |
|  | 100K+ | 13.5% |
| *Education* | Some high school or less | 0.0% |
|  | High school diploma or G.E.D. | 6.2% |
|  | Some college | 17.6% |
|  | Degree from a vocational school | 7.7% |
|  | Undergraduate degree | 41.1% |
|  | Graduate degree | 27.4% |
| *Employment* | Working full-time | 76.6% |
|  | Working part-time (at least 25 hours per week) | 6.6% |
|  | A homemaker | 1.7% |
|  | Not currently working and not seeking employment | 2.5% |
|  | Not currently working, but seeking employment | 4.1% |
|  | A full-time student | 0.8% |
|  | A part-time student | 0.0% |
|  | Retired | 1.7% |
|  | Working full-time under furlough | 4.8% |
|  | Working part-time under furlough | 1.2% |
| *Area* | Rural | 23.7% |
|  | Suburban | 43.8% |
|  | Urban | 32.6% |
| *Political Orientation* | Very Liberal | 14.7% |
|  | Liberal or Slightly Liberal | 33.0% |
|  | Conservative or Slightly Conservative | 40.7% |
|  | Very Conservative | 11.6% |

**Appendix A2a:** All Risk Taking by Stepwise Regression (Demographic, Situational, Dispositional)

| **Model** |  | **Unstandardized** | **Standard Error** | **Standardized** | **t** | **p** |
| --- | --- | --- | --- | --- | --- | --- |
| H<sub>0</sub> | (Intercept) | 3.472 | 0.070 |  | 49.622 | < .001 |
| H<sub>1</sub> | (Intercept) | 3.884 | 1.234 |  | 3.147 | 0.002 |
|  | Employ | -0.028 | 0.014 | -0.089 | -1.980 | 0.048 |
|  | Income | -0.134 | 0.055 | -0.108 | -2.453 | 0.015 |
|  | Education | 0.175 | 0.057 | 0.140 | 3.072 | 0.002 |
|  | Age | -0.013 | 0.006 | -0.099 | -2.267 | 0.024 |
|  | Race | 0.018 | 0.062 | 0.012 | 0.289 | 0.773 |
|  | Parent | -0.402 | 0.138 | -0.131 | -2.904 | 0.004 |
|  | Older than 60 | -0.150 | 0.147 | -0.044 | -1.017 | 0.310 |
|  | Urban - Rural | -0.091 | 0.088 | -0.044 | -1.028 | 0.304 |
|  | Political Orientation | 0.246 | 0.041 | 0.260 | 6.006 | < .001 |
|  | Gender | 0.344 | 0.124 | 0.117 | 2.783 | 0.006 |

**Appendix A2b:**
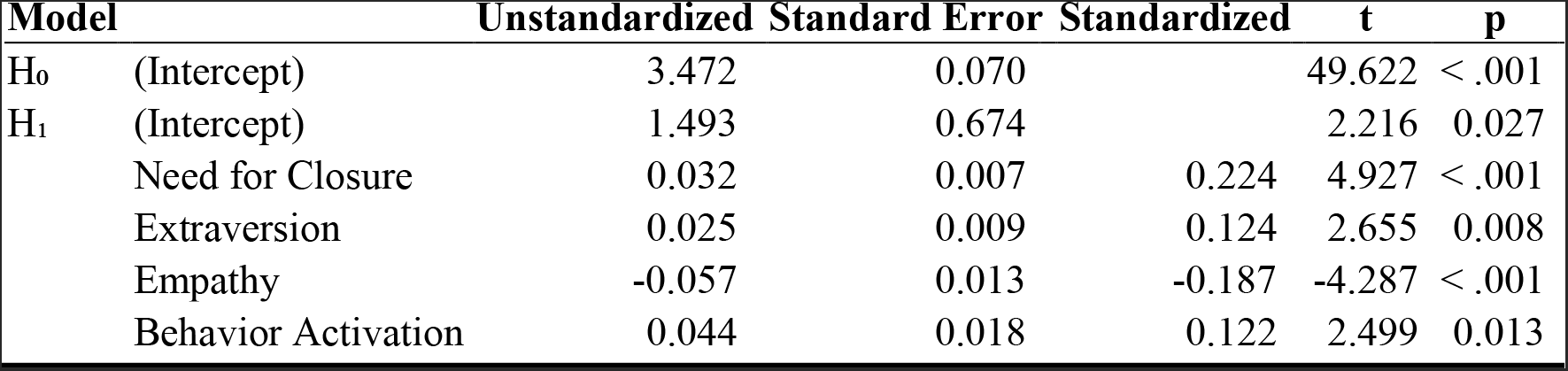
All Risk Taking by Sig. Dispositional Variables

**Appendix A2c:** All Risk Taking by Dispositional + Situational

| Model |  | Unstandardized | Standard Error | Standardized | t | p |
| --- | --- | --- | --- | --- | --- | --- |
| H<sub>0</sub> | (Intercept) | 3.118 | 0.367 |  | 8.489 | < .001 |
|  | Living Space Size | 0.030 | 0.068 | 0.020 | 0.438 | 0.662 |
|  | Perceived Scarcity | 0.184 | 0.041 | 0.203 | 4.502 | < .001 |
|  | Age | -0.014 | 0.006 | -0.106 | -2.326 | 0.020 |
| H<sub>1</sub> | (Intercept) | 1.669 | 0.739 |  | 2.258 | 0.024 |
|  | Living Space Size | 0.035 | 0.066 | 0.024 | 0.529 | 0.597 |
|  | Perceived Scarcity | 0.101 | 0.043 | 0.111 | 2.356 | 0.019 |
|  | Age | -0.012 | 0.006 | -0.086 | -1.948 | 0.052 |
|  | Need for Closure | 0.026 | 0.007 | 0.183 | 3.832 | < .001 |
|  | Extraversion | 0.022 | 0.009 | 0.111 | 2.362 | 0.019 |
|  | Behavior Activation | 0.042 | 0.018 | 0.117 | 2.403 | 0.017 |
|  | Empathy | -0.050 | 0.013 | -0.164 | -3.715 | < .001 |

**Appendix A3a:** Nonparametric Regressions of High Risk Taking by Dispositional + Situational Clusters

| Model |  | Unstandardized | Standard Error | Standardized | t | p |
| --- | --- | --- | --- | --- | --- | --- |
| H<sub>0</sub> | (Intercept) | 2.341 | 0.438 |  | 5.339 | < .001 |
|  | Living Space Size | -0.205 | 0.081 | -0.110 | -2.541 | 0.011 |
|  | Perceived Scarcity | 0.381 | 0.049 | 0.332 | 7.830 | < .001 |
|  | Age | -0.024 | 0.007 | -0.140 | -3.277 | 0.001 |
| H<sub>1</sub> | (Intercept) | -0.285 | 0.912 |  | -0.312 | 0.755 |
|  | Living Space Size | -0.218 | 0.076 | -0.117 | -2.861 | 0.004 |
|  | Perceived Scarcity | 0.243 | 0.050 | 0.212 | 4.881 | < .001 |
|  | Age | -0.023 | 0.007 | -0.132 | -3.266 | 0.001 |
|  | Extraversion | 0.054 | 0.011 | 0.211 | 4.977 | < .001 |
|  | Need for Closure | 0.032 | 0.008 | 0.175 | 4.139 | < .001 |
|  | Behavior Activation | 0.051 | 0.012 | 0.184 | 4.312 | < .001 |
|  | Empathy | -0.068 | 0.016 | -0.177 | -4.335 | < .001 |

**Appendix A3b:**
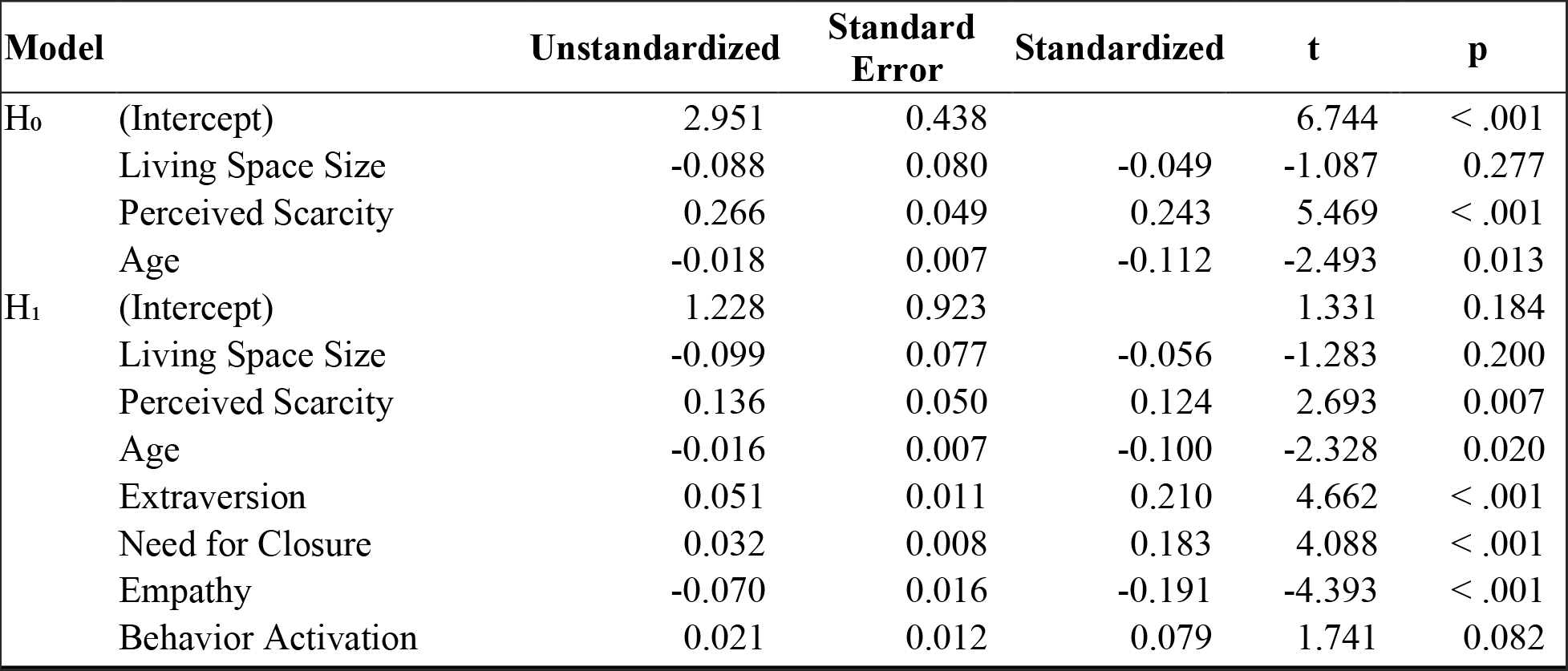
Nonparametric Regressions of Low Risk Taking by Dispositional + Situational Clusters

**Appendix 34c:** Regressions of Essential Activity by Dispositional + Situational Clusters

| Model |  | Unstandardized | Standard Error | Standardized | t | p |
| --- | --- | --- | --- | --- | --- | --- |
| H<sub>0</sub> | (Intercept) | 3.285 | 0.380 |  | 8.651 | < .001 |
|  | Living Space Size | 0.147 | 0.070 | 0.099 | 2.100 | 0.036 |
|  | Perceived Scarcity | 0.102 | 0.042 | 0.110 | 2.408 | 0.016 |
|  | Age | -0.010 | 0.006 | -0.075 | -1.627 | 0.104 |
| H<sub>1</sub> | (Intercept) | 2.556 | 0.835 |  | 3.061 | 0.002 |
|  | Living Space Size | 0.142 | 0.070 | 0.095 | 2.036 | 0.042 |
|  | Perceived Scarcity | 0.048 | 0.046 | 0.053 | 1.061 | 0.289 |
|  | Age | -0.010 | 0.006 | -0.069 | -1.501 | 0.134 |
|  | Extraversion | 0.019 | 0.010 | 0.094 | 1.942 | 0.053 |
|  | Need for Closure | 0.014 | 0.007 | 0.094 | 1.951 | 0.052 |
|  | Behavior Activation | 0.008 | 0.011 | 0.037 | 0.763 | 0.446 |
|  | Empathy | -0.027 | 0.014 | -0.088 | -1.891 | 0.059 |

**Table A4.**
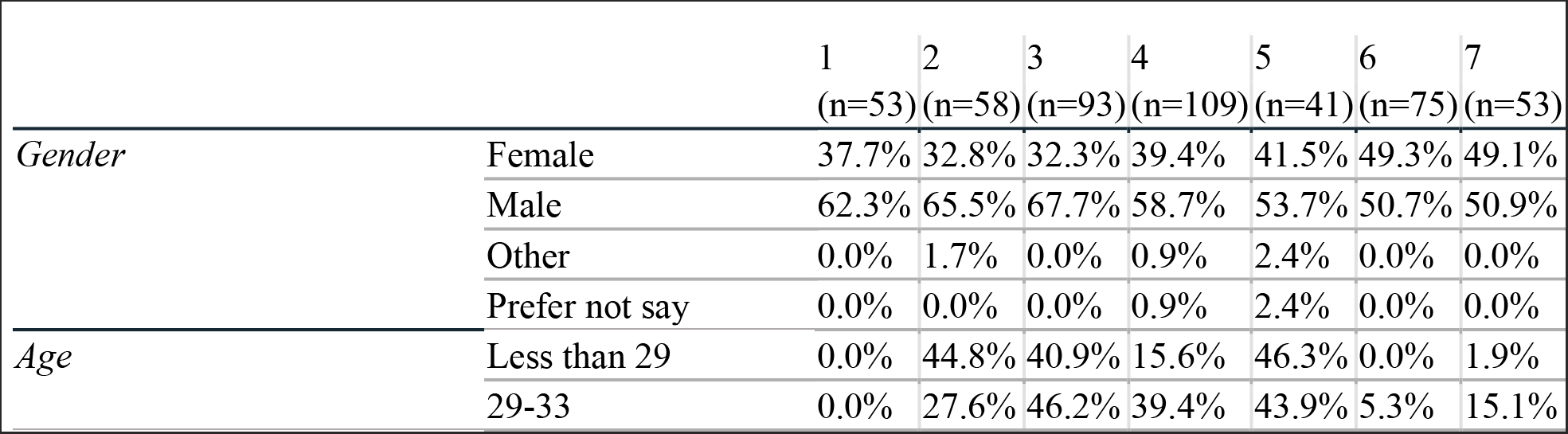
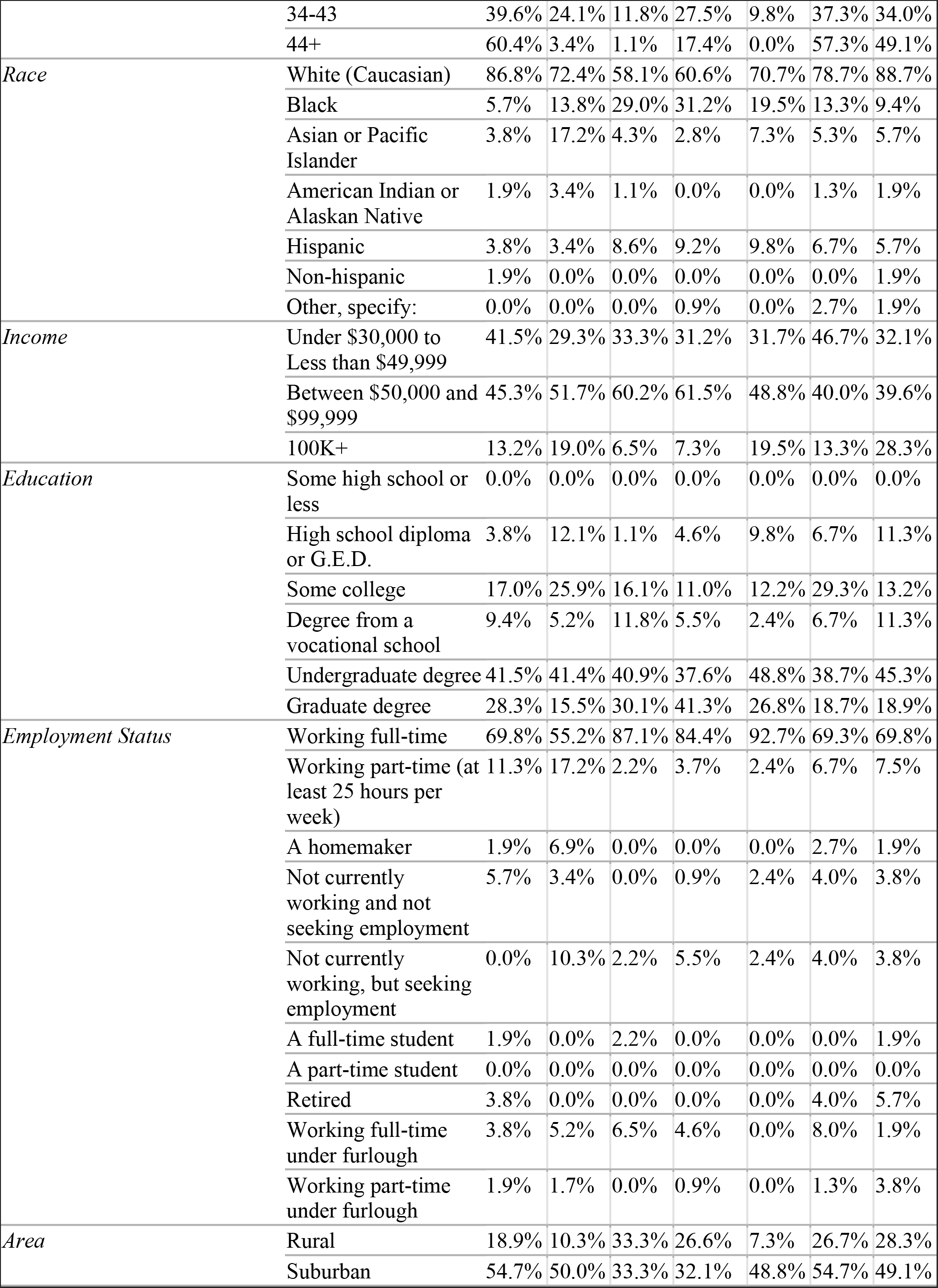
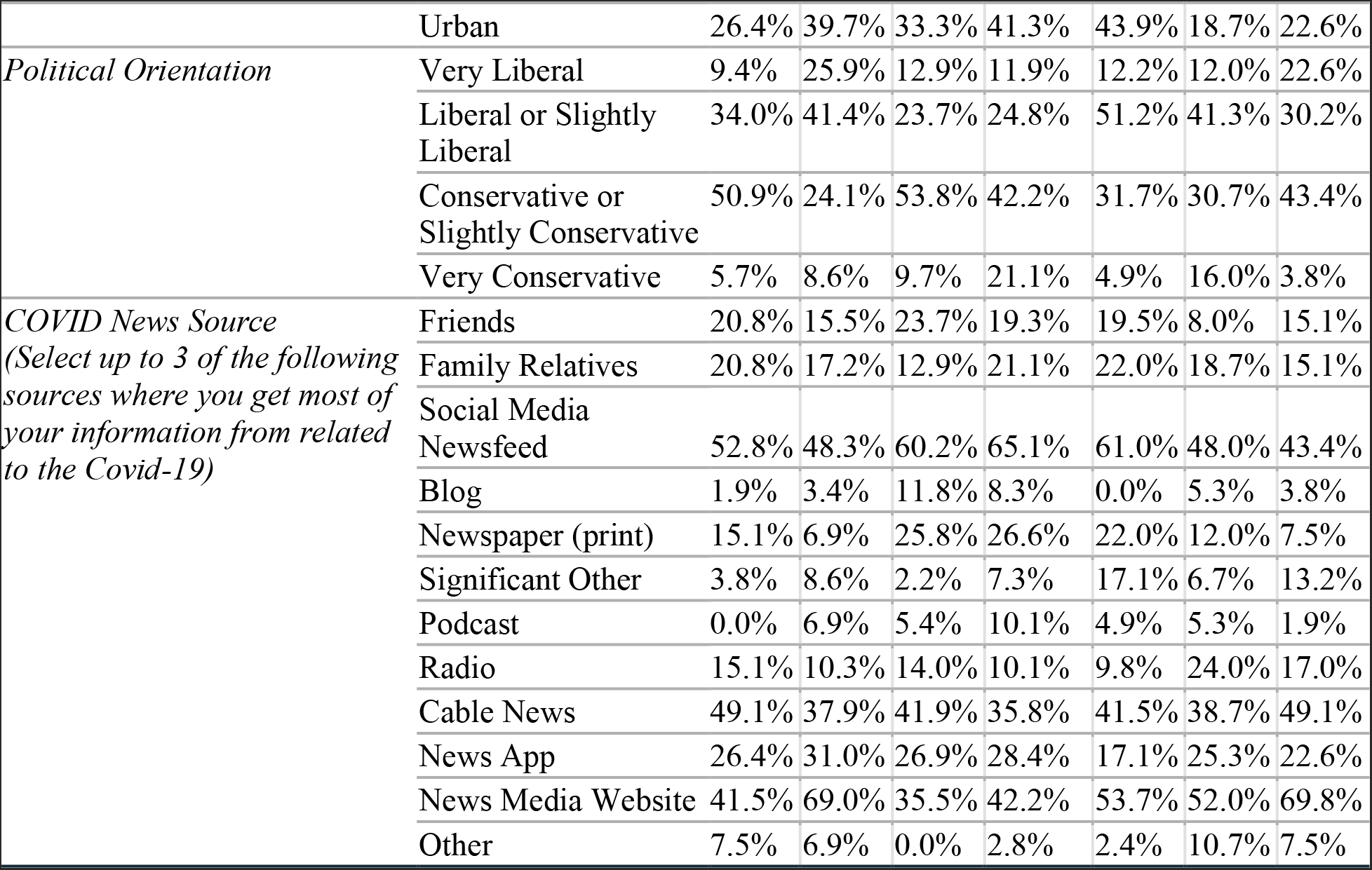
Demographic breakdown of Cluster Groups (column percentages)

**Table A5.**
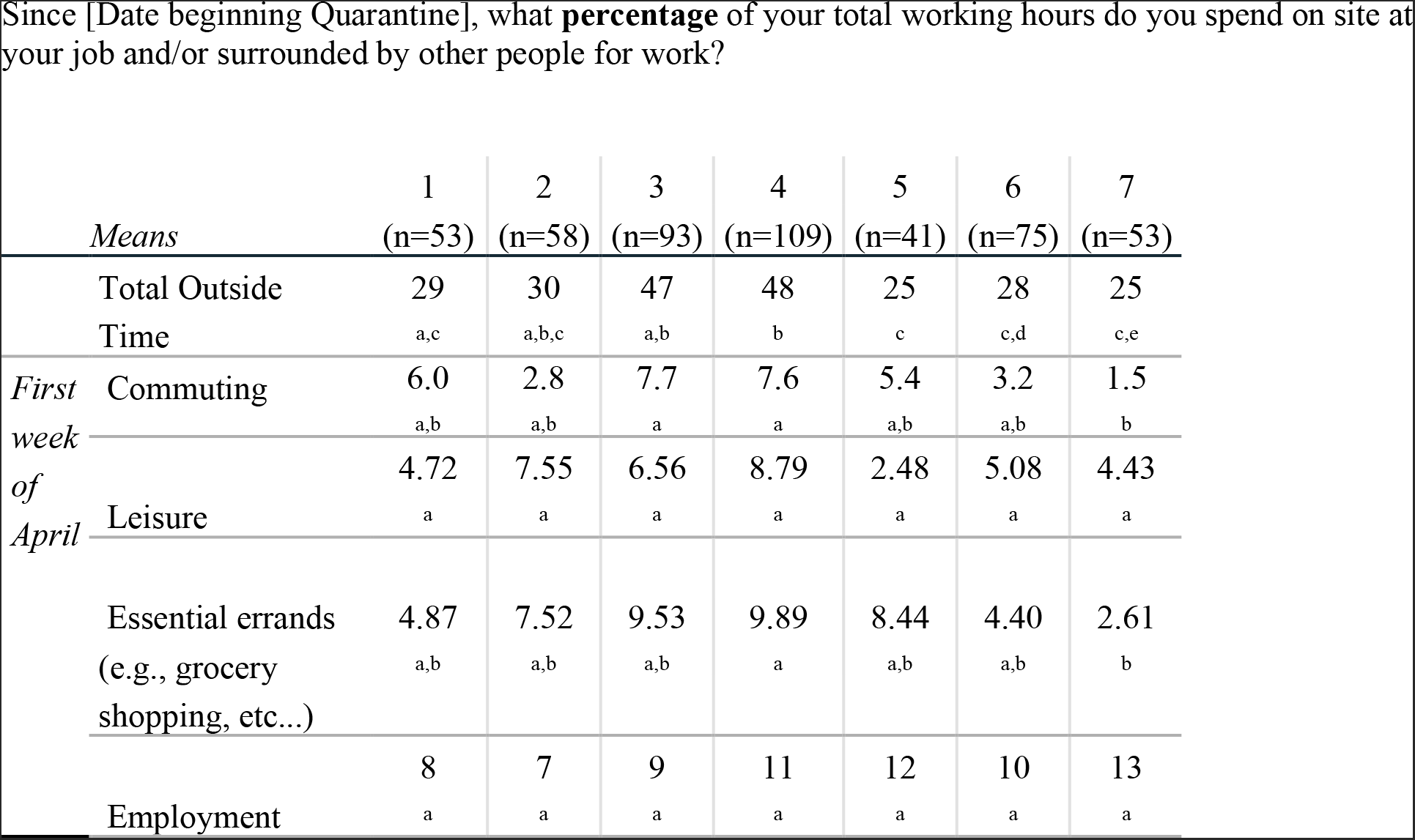
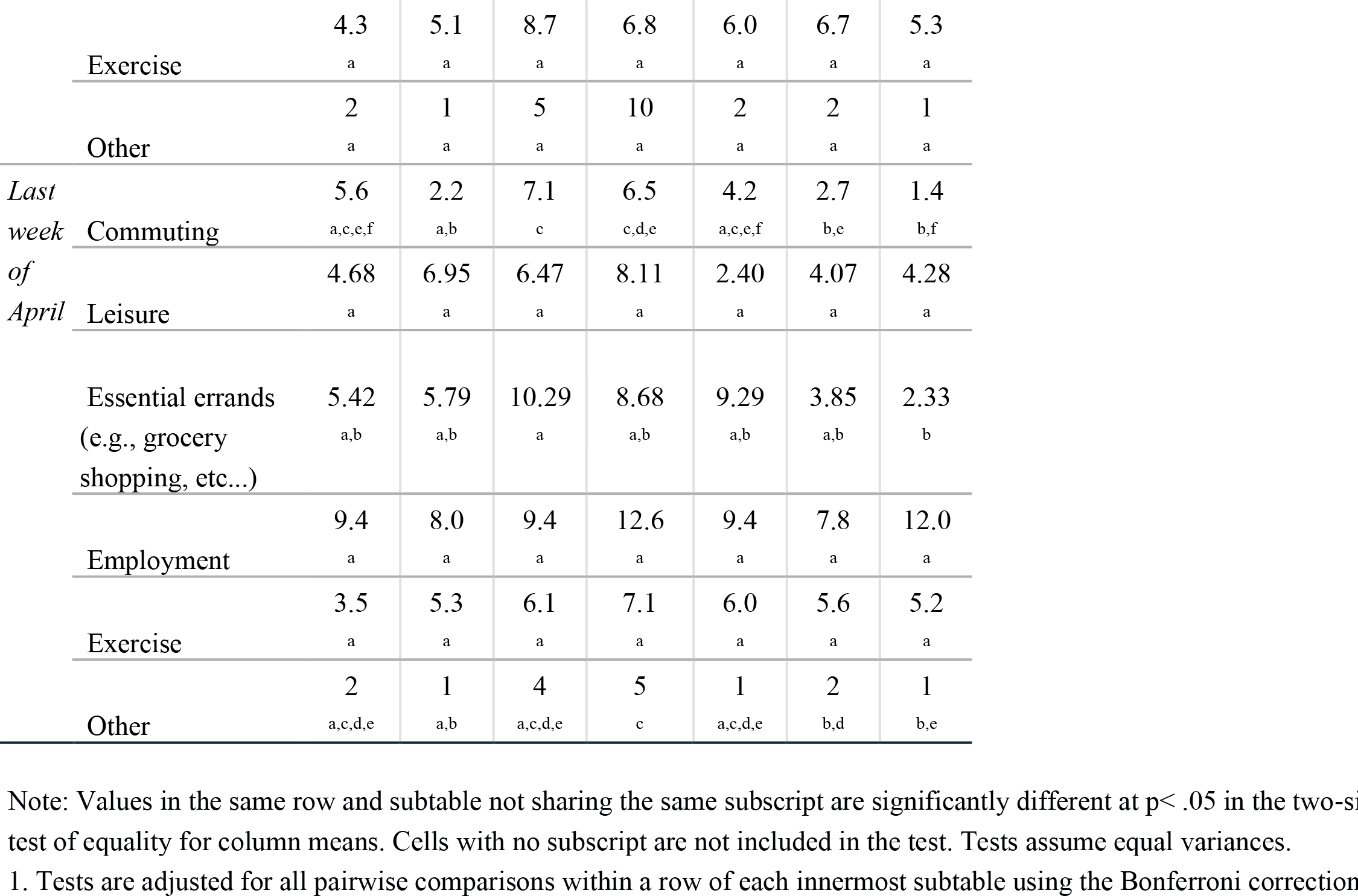
Hours outside by Cluster Groups (mean comparisons)

**Table A6.**
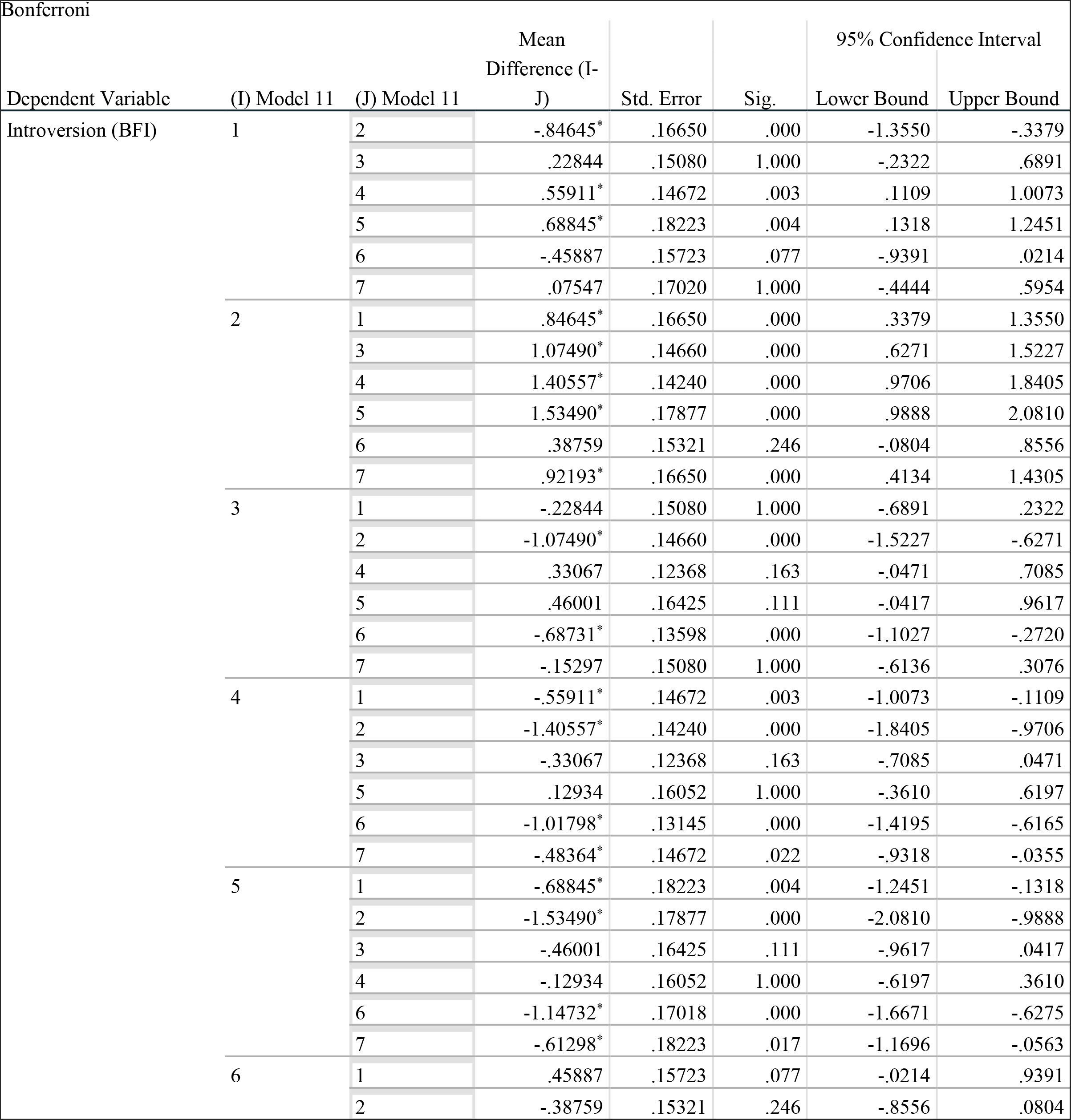
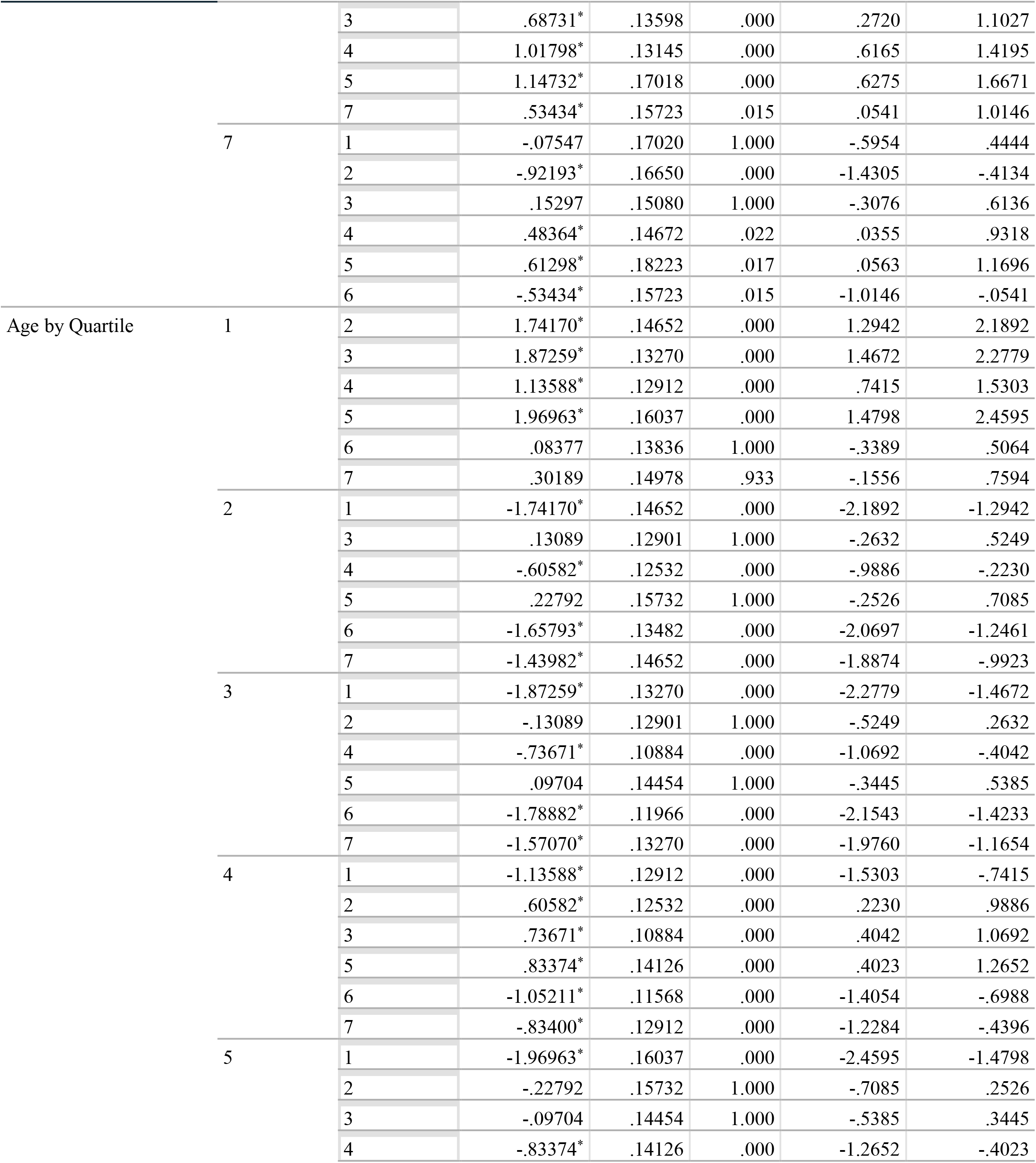
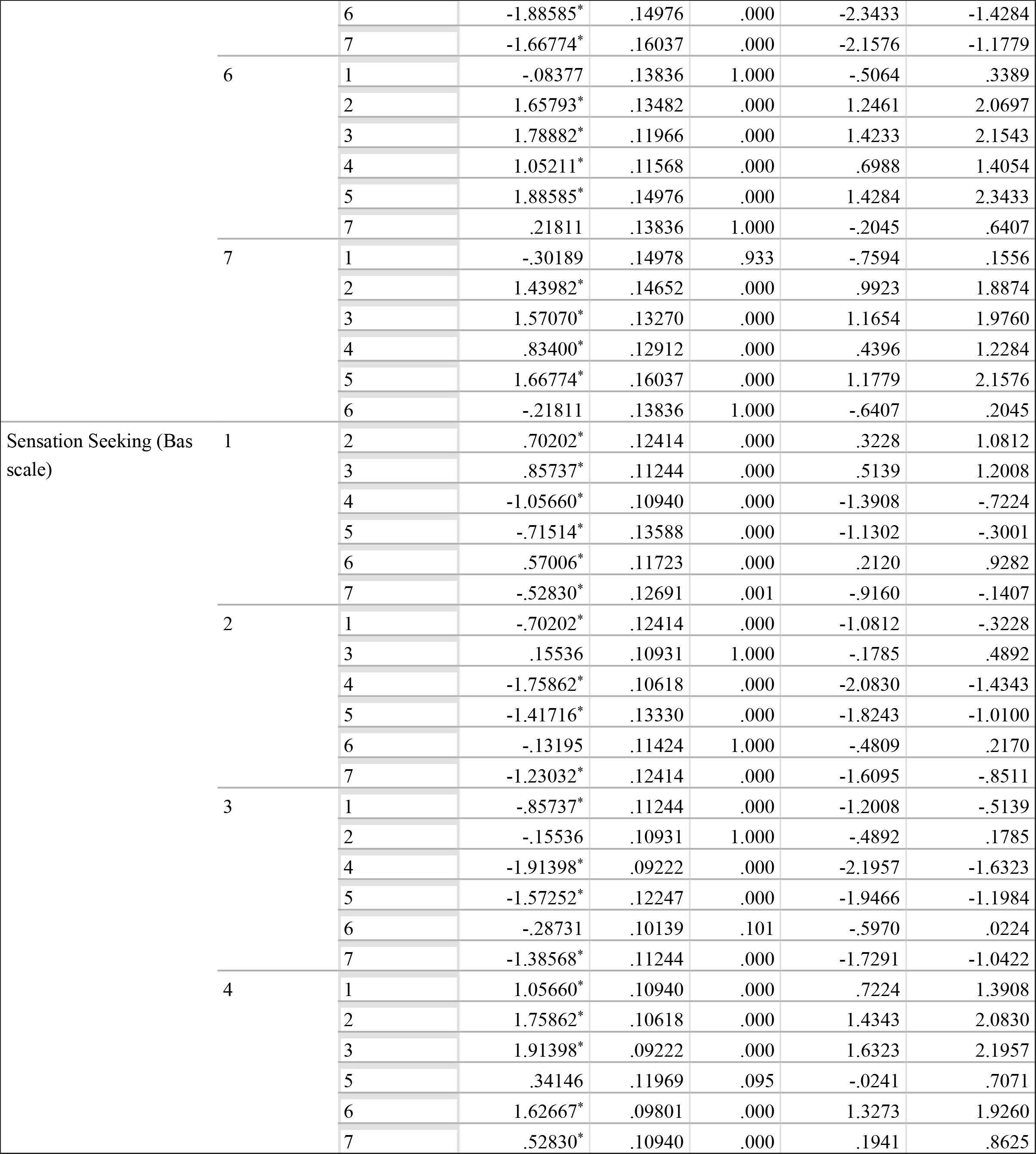
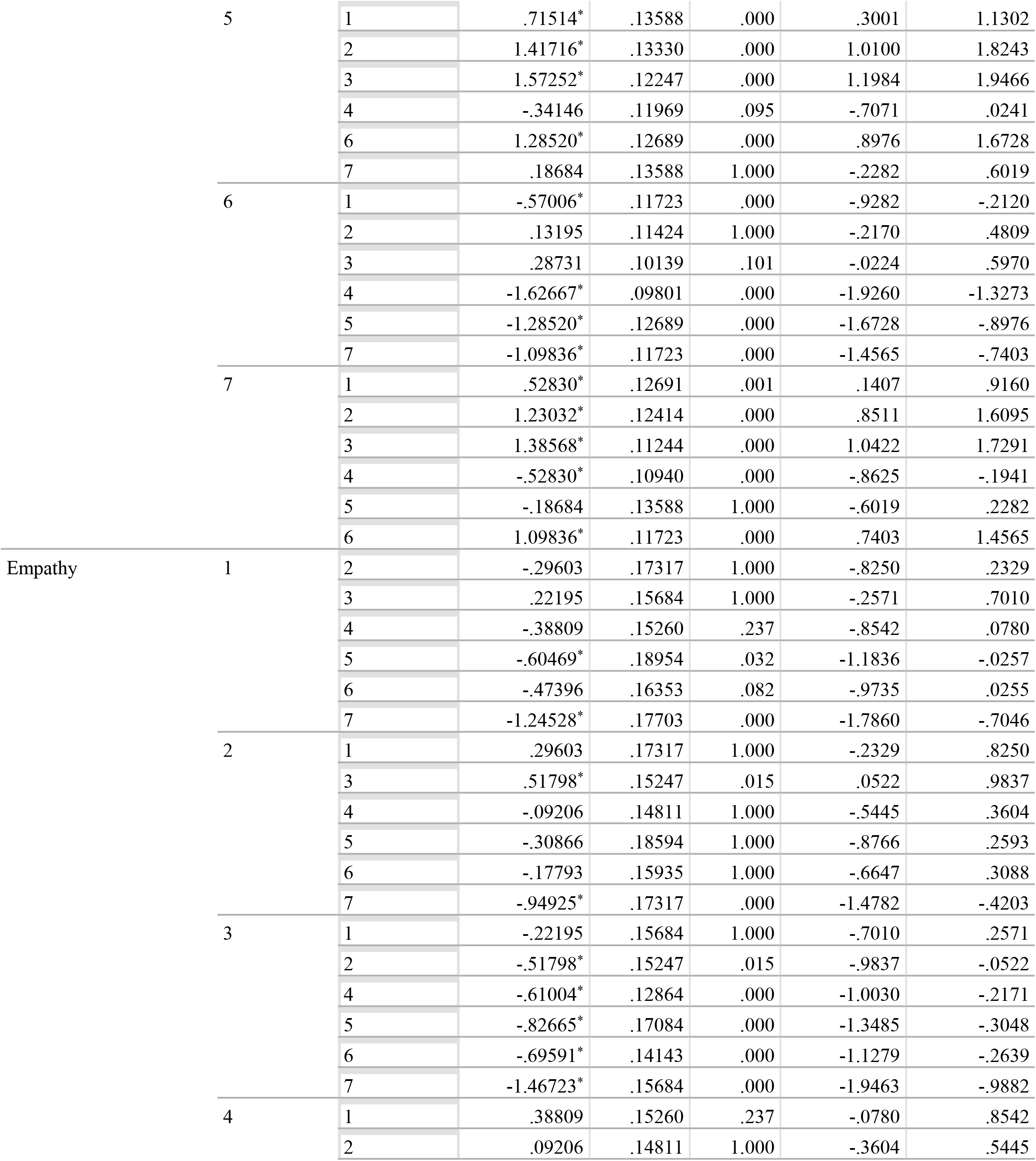
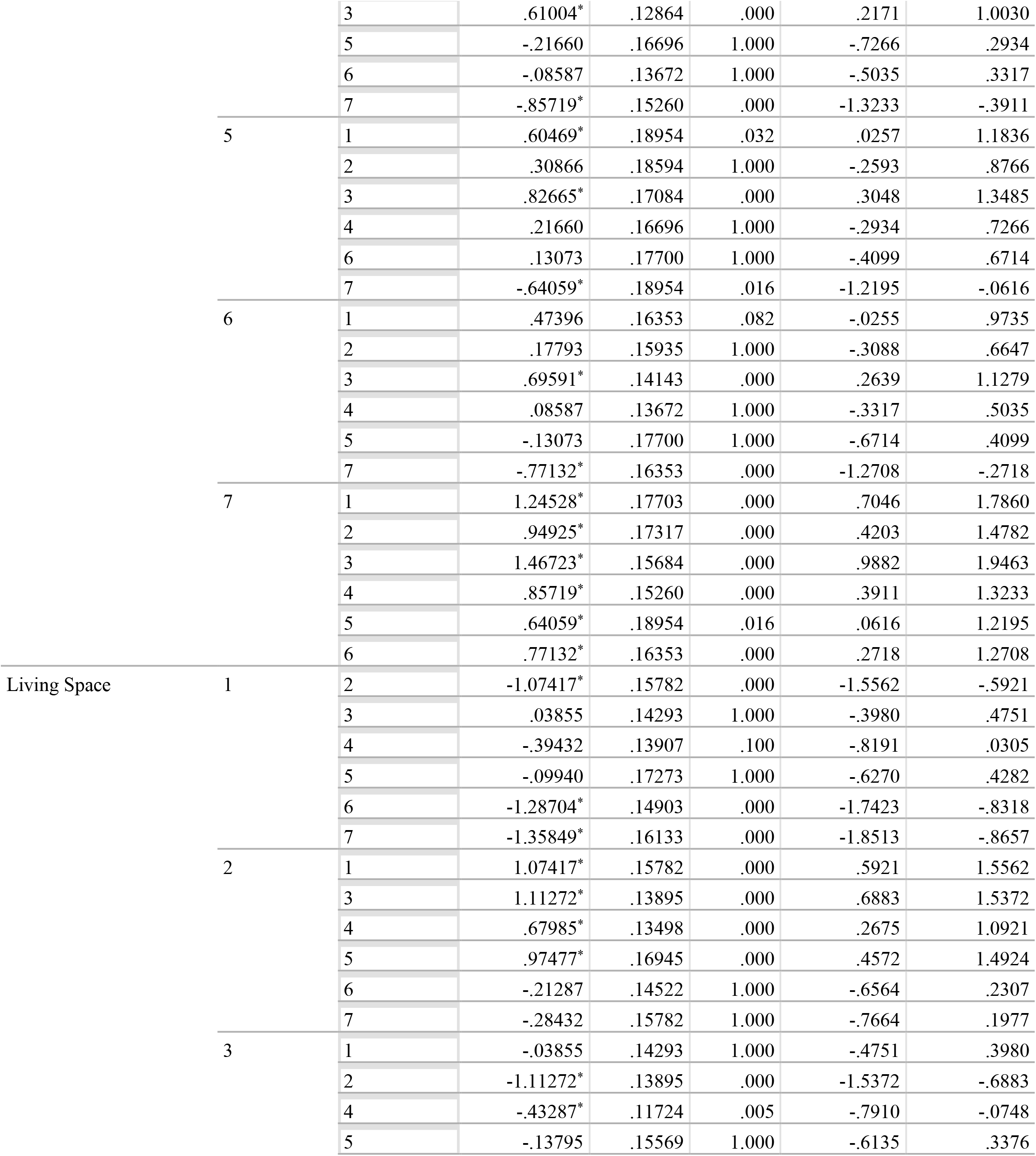
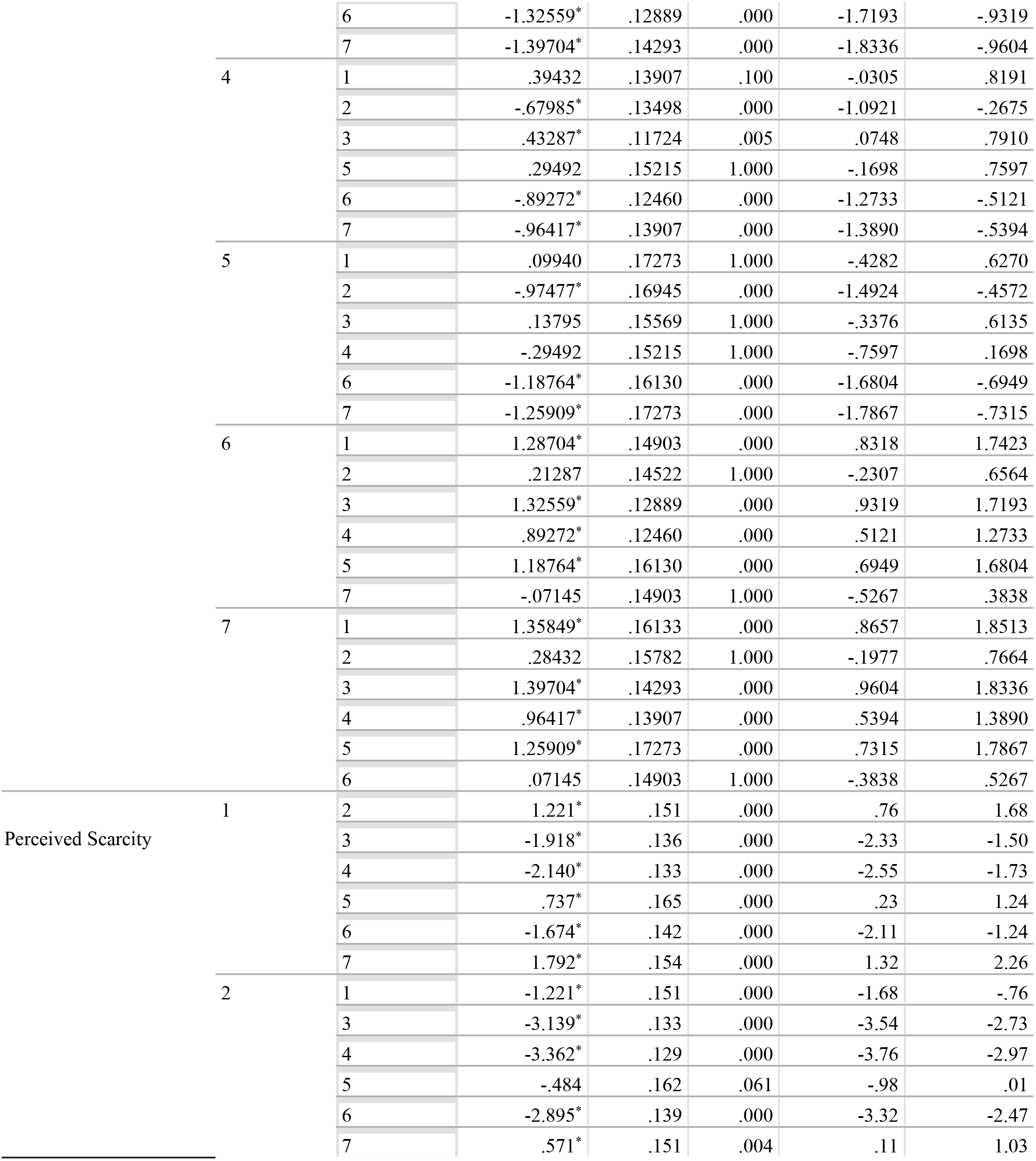
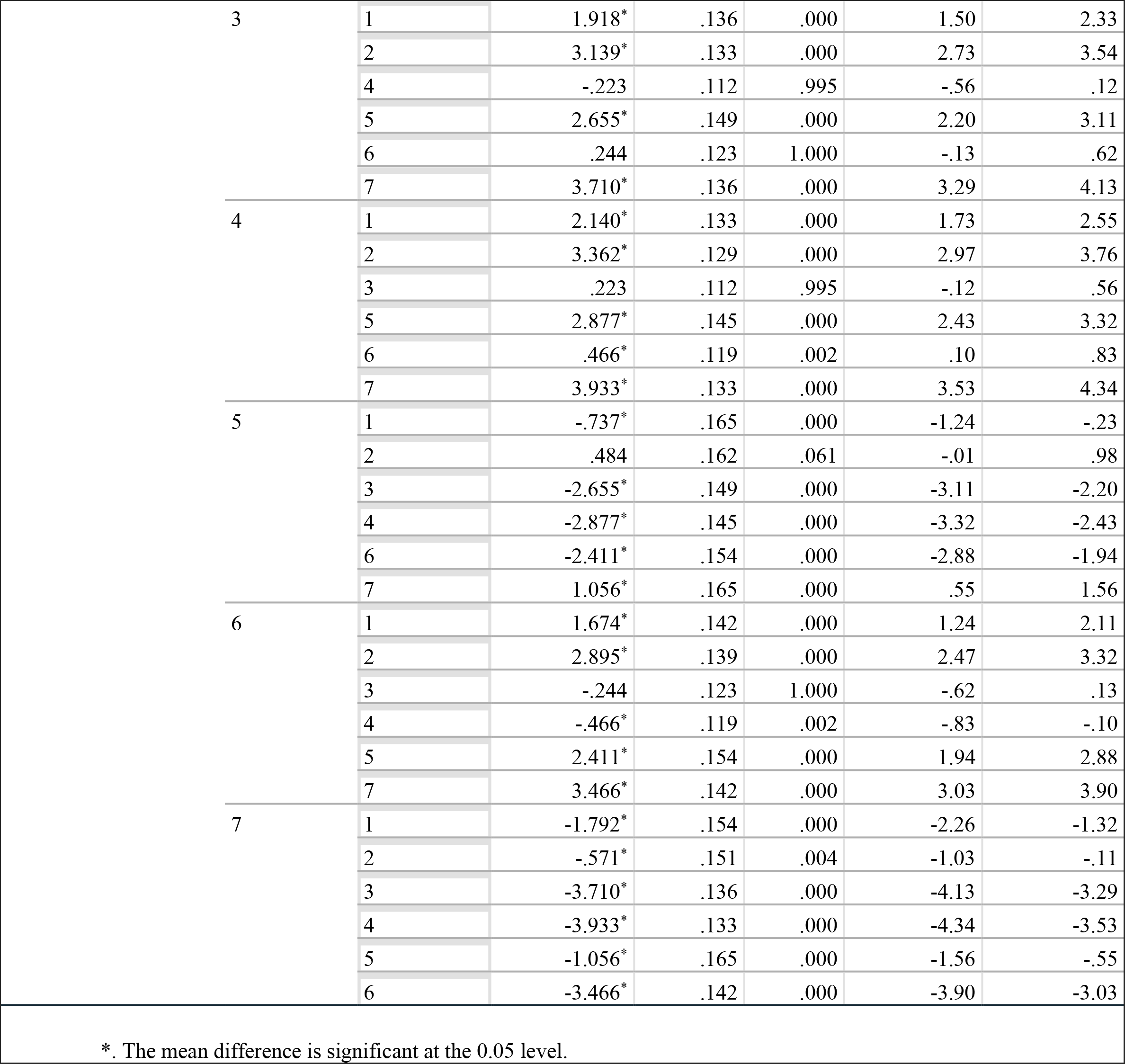
Multiple Comparisons between Cluster groups by input variables

**Table A6b.** ANOVA Results of Input Variables from Cluster Model

|  |  | ANOVA |  |  |  |  |
| --- | --- | --- | --- | --- | --- | --- |
|  |  | Sum of Squares | df | Mean Square | F | Sig. |
| Introversion (BFI) | Between Groups | 114.028 | 6 | 19.005 | 24.756 | .000 |
|  | Within Groups | 364.652 | 475 | .768 |  |  |
|  | Total | 478.680 | 481 |  |  |  |
| Age by Quartile | Between Groups | 284.238 | 6 | 47.373 | 79.683 | .000 |
|  | Within Groups | 282.395 | 475 | .595 |  |  |
|  | Total | 566.633 | 481 |  |  |  |
| Sensation-Seeking (BisBas) | Between Groups | 278.591 | 6 | 46.432 | 108.787 | .000 |
|  | Within Groups | 202.736 | 475 | .427 |  |  |
|  | Total | 481.328 | 481 |  |  |  |
| Empathy (IRI) | Between Groups | 83.510 | 6 | 13.918 | 16.760 | .000 |
|  | Within Groups | 394.473 | 475 | .830 |  |  |
|  | Total | 477.983 | 481 |  |  |  |
| Living Space | Between Groups | 150.359 | 6 | 25.060 | 36.333 | .000 |
|  | Within Groups | 327.624 | 475 | .690 |  |  |
|  | Total | 477.983 | 481 |  |  |  |
| Perceived Scarcity | Between Groups | 1089.129 | 6 | 181.522 | 288.639 | .000 |
|  | Within Groups | 298.722 | 475 | .629 |  |  |
|  | Total | 1387.851 | 481 |  |  |  |

Risk Behavior by Factor and Cluster

**Risk Behavior by Factor and Cluster**
| CRI Factor. Low Risk |  |  | High Risk |  |
| --- | --- | --- | --- | --- |
| Cluster | Mean | SD | Mean | SD |
| 1 | 2.918 | 1.598 | 2.057 | 1.528 |
| 2 | 2.529 | 1.653 | 1.750 | 1.554 |
| 3 | 3.842 | 1.966 | 3.602 | 2.144 |
| 4 | 3.599 | 2.035 | 2.844 | 2.247 ** |
| 5 | 2.846 | 1.616 | 1.756 | 1.351 ** |
| 6 | 2.556 | 1.663 | 1.920 | 1.681 ** |
| 7 | 2.547 | 1.539 | 1.236 | 0.812 * |

## The Covid Risk-taking Inventory

The Covid Risk-taking Inventory is an adaptation of the Benthin Risk Perception Scale (Benthin et al., 1993) to reflect the guidelines set forth by the Centers for Disease Control and Prevention (CDC) that were updated on April 13th, 2020, advising the public on precautionary and personal hygiene habits to mitigate the spread of COVID-19. We created scenarios based on the 10 recommendations and risk scenarios sent to United States businesses.

### Overview

The following ten items were developed to assess risk perception during the COVID-19 pandemic. Each participant received a general prompt to think about how their lives had changed in the last few weeks and then to consider the following activities. Every activity item then had four questions asking about the following: (1) frequency of engagement since the estimated date when they first began modifying their behavior due to the coronavirus, (2) a risk-to-benefit or risk-to-‘necessity’ evaluation, and (3) risk assessment of consequences from the activity relative to the self and (4) consequences from the activity relative to others.

| **<u>Activity Items</u>** | **<u>Risk Evaluation</u>** | **<u>Factor Loading</u>** |
| --- | --- | --- |
| 1. Going shopping at the grocery store | **<u>Essential</u>** | **0.621** |
| 2. Being outside without a face mask | **<u>Essential</u>** | **0.518** |
| 3. Meeting a friend in person while maintaining “social distancing” guidelines | **<u>Low Risk</u>** | **0.476** |
| 4. Returning home from a store without washing your hands | **<u>Low Risk</u>** | **0.679** |
| 5. Arranging an in-person meeting with someone you do not know well for essential business | **<u>High Risk</u>** | **0.680** |
| 6. Visiting a public space (e.g. beach, park) | **<u>Low Risk</u>** | **0.614** |
| 7. Using a car share service (such as Uber or Lyft) or taxi | **<u>Travel</u>**<br>(not included) | **0.860** |
| 8. Using public transit while taking appropriate precautions | **<u>Travel</u>**<br>(not included) | **0.644** |
| 9. Exercising outside alone while adhering to “social distancing” guidelines | **<u>Essential</u>** | **0.821** |
| 10. Attending a private event or celebration with more than five other guests from outside of your household | **<u>High Risk</u>** | **0.822** |

## The Covid Risk-Taking Inventory

### Questions for Each Activity Item

#### Risk-taking behavior

1) How many times have you engaged in this activity since …

a. Never
b. Once or twice in the past 2 months
c. Once or twice in the past month
d. About once a week
e. A few days a week
f. Just about every day
g. Every single day
h. Multiple times a day

#### Risk-to-benefits Evaluation

2) How would you compare the benefits or necessity of this activity with the risks?

a. Risks are much greater than the benefits
b. Risks are somewhat greater than benefits
c. Benefits are somewhat greater than risks
d. Benefits are much greater than risks

#### Personal Risk Assessment

3) If you did this activity, how much are you at risk for something bad happening?

a. High risk
b. Moderate risk
c. Low risk
d. No risk at all

#### Interpersonal Risk Assessment

4) If something bad happened to someone else because of this activity, how serious would it be?

a. Not at all serious
b. A little serious
c. Pretty serious
d. Very serious

## References

1. Allen-Ebrahimian B. Timeline: The early days of China’s coronavirus outbreak and cover-up [Internet]. Axios. 2020 [cited 2020 May 11]. Available from: https://www.axios.com/timeline-the-early-days-of-chinascoronavirus-outbreak-and-cover-up-ee65211a-afb6-4641-97b8-353718a5faab.html

2. Heller K. Here’s what major cities look like now that the coronavirus has shut everything down [Internet]. Washington Post. 2020 [cited 2020 May 17]. Available from: https://www.washingtonpost.com/graphics/2020/national/coronavirus-shutdowns-cities/

3. Biron B. Experts green light online shopping amid the coronavirus – Business Insider [Internet]. 2020 [cited 2020 May 17]. Available from: https://www.businessinsider.com/experts-green-light-online-shoppingamid-the-coronavirus-2020-3

4. Dwyer C, Aubrey A. CDC Now Recommends Americans Consider Wearing Cloth Face Coverings In Public [Internet]. NPR. 2020 [cited 2020 May 17]. Available from: https://www.npr.org/sections/coronaviruslive-updates/2020/04/03/826219824/president-trump-says-cdc-now-recommends-americans-wear-clothmasks-in-public

5. Mervosh S, Lu D, Swales V. See Which States and Cities Have Told Residents to Stay at Home. The New York Times [Internet]. 2020 Mar 31 [cited 2020 May 17]; Available from: https://www.nytimes.com/interactive/2020/us/coronavirus-stay-at-home-order.html

6. Stay Home Save Lives [Internet]. 2020 [cited 2020 May 11]. Available from: https://govstatus.egov.com/stayhomesavelives

8. 16 national health care leaders. The best thing everyday Americans can do to fight coronavirus? #StayHome, save lives [Internet]. USA TODAY. 2020 [cited 2020 May 11]. Available from: https://www.usatoday.com/story/opinion/2020/03/15/coronavirus-stay-home-hel-america-save-livescolumn/5054241002/

8. Marcus J. Quarantine Fatigue Is Real [Internet]. The Atlantic. 2020 [cited 2020 May 11]. Available from: https://www.theatlantic.com/ideas/archive/2020/05/quarantine-fatigue-real-and-shaming-people-wonthelp/611482/

9. CDC. Coronavirus Disease 2019 (COVID-19) [Internet]. Centers for Disease Control and Prevention. 2020 [cited 2020 May 11]. Available from: https://www.cdc.gov/coronavirus/2019-ncov/prevent-gettingsick/social-distancing.html

10. Mays J.C, Newman A. Virus Is Twice as Deadly for Black and Latino People Than Whites in N.Y.C. The New York Times [Internet]. 2020 May 7 [cited 2020 May 10]; Available from: https://www.nytimes.com/2020/04/08/nyregion/coronavirus-race-deaths.html

11. The Editorial Board. Opinion | ‘You’re On Your Own,’ Essential Workers Are Being Told. The New York Times [Internet]. 2020 Apr 20 [cited 2020 May 17]; Available from: https://www.nytimes.com/2020/04/20/opinion/osha-coronavirus.html

12. Gould E. Lack of paid sick days and large numbers of uninsured increase risks of spreading the coronavirus [Internet]. Economic Policy Institute. 2020 [cited 2020 May 11]. Available from: https://www.epi.org/blog/lack-of-paid-sick-days-and-large-numbers-of-uninsured-increase-risks-ofspreading-the-coronavirus/

13. Weintraub K. Lack of Paid Sick Leave a Coronavirus Threat [Internet]. WebMD. 2020 [cited 2020 May 11]. Available from: https://www.webmd.com/lung/news/20200310/lack-of-paid-sick-leave-a-coronavirusthreat

14. Tian F, Li H, Tian S, Yang J, Shao J, Tian C. Psychological symptoms of ordinary Chinese citizens based on SCL-90 during the level I emergency response to COVID-19. Psychiatry Research. 2020 Apr 11: 112992.

15. Cao, W., Fang, Z., Hou, G., Han, M., Xu, X., Dong, J., & Zheng, J. (2020). The psychological impact of the COVID-19 epidemic on college students in China. Psychiatry research, 112934.

16. Flesia L, Fietta V, Colicino E, Segatto B, Monaro M. Stable psychological traits predict perceived stress related to the COVID-19 outbreak. 2020. doi:10.31234/osf.io/yb2h8.

17. Serafim AP, Gonçalves PD, Rocca CC, Lotufo Neto F. The impact of COVID-19 on Brazilian mental health through vicarious traumatization[published online ahead of print, 2020 May 11]. Braz J Psychiatry. 2020;S1516–44462020005013204. doi:10.1590/1516-4446–2020-0999

18. Chew C, Eysenbach G. Pandemics in the Age of Twitter: Content Analysis of Tweets during the 2009 H1N1 Outbreak. PLOS ONE. 2010 Nov 29;5(11): e14118.

19. Cava MA, Fay KE, Beanlands HJ, McCay EA, Wignall R. Risk perception and compliance with quarantine during the SARS outbreak. Journal of Nursing Scholarship. 2005 Dec;37(4):343–7.

20. Arndt J, Schimel J, Goldenberg JL. Death can be good for your health: Fitness intentions as a proximal and distal defense against mortality salience 1. Journal of Applied Social Psychology. 2003 Aug;33(8):1726–46.

21. Västfjäll D, Peters E, Slovic P. The affect heuristic, mortality salience, and risk: Domain‐specific effects of a natural disaster on risk‐benefit perception. Scandinavian journal of psychology. 2014 Dec;55(6):527–32.

22. Taubman–Ben-Ari, O., & Skvirsky, V. (2019). The Terror Management Underpinnings of Risky Behavior. In Handbook of Terror Management Theory (pp. 559–576). Academic Press.

23. Miller RL, Mulligan RD. Terror management: The effects of mortality salience and locus of control on risk-taking behaviors. Personality and Individual differences. 2002 Nov 1;33(7):1203–14.

24. Hanoch Y, Rolison JJ, Freund AM. Does medical risk perception and risk taking change with age?. Risk Analysis. 2018 May;38(5):917–28.

25. Schumpe BM, Brizi A, Giacomantonio M, Panno A, Kopetz C, Kosta M, et al. Need for Cognitive Closure decreases risk taking and motivates discounting of delayed rewards. Personality and Individual Differences. 2017 Mar, 1; 107:66–71.

26. Galloway G, Lopez K. Sensation seeking and attitudes to aspects of national parks: a preliminary empirical investigation. Tourism Management. 1999 Dec 1;20(6):665–71.

27. Cheng ASK Lee HC. Risk-taking behavior and response inhibition of commuter motorcyclists with different levels of impulsivity. Transportation Research Part F: Traffic Psychology and Behaviour. 2012 Sep 1;15(5):535–43.

28. Boney-McCoy S, Gibbons FX, Gerrard M. Self-Esteem, Compensatory Self-Enhancement, and the Consideration of Health Risk. Pers Soc Psychol Bull. 1999 Aug 1;25(8):954–65.

29. Neuberg SL, Kenrick DT, Schaller M. Human Threat Management Systems: Self-Protection and Disease Avoidance. Neurosci Biobehav Rev. 2011 Mar;35(4):1042–51.

30. Carpentier MY, Mullins LL, Elkin TD, Wolfe‐Christensen C. Predictors of health‐harming and health‐protective behaviors in adolescents with cancer. Pediatric blood & cancer. 2008 Oct;51(4):525–30.

31. Zurcher A. Trump defends tweets against US states’ lockdowns. BBC News [Internet]. 2020 Apr 18 [cited 2020 May 11]; Available from: https://www.bbc.com/news/world-us-canada-52330531

32. Sanders L. Yahoo News/YouGov: When America believes the country will be ready to reopen | YouGov [Internet]. 2020 [cited 2020 May 11]. Available from: https://today.yougov.com/topics/politics/articles-reports/2020/04/20/when-america-reopens-poll

33. Levin S, Ho V. Thousands of people pack California beaches despite coronavirus concerns [Internet]. the Guardian. 2020 [cited 2020 May 11]. Available from: http://www.theguardian.com/usnews/2020/apr/27/california-beaches-coronavirus-orange-county

34. Maryland Transportation Institute (2020). University of Maryland COVID-19 Impact Analysis Platform. University of Maryland, College Park, USA [Internet]. 2020 [cited 2020 May 11]; Available from: https://data.covid.umd.edu

35. Anderson CA, Allen JJ, Plante C, Quigley-McBride A, Lovett A, Rokkum JN. The MTurkification of Social and Personality Psychology. Pers Soc Psychol Bull. 2019 Jun 1;45(6):842–50.

36. U.S. Census Bureau. U.S. Census Bureau QuickFacts: United States [Internet]. [cited 2020 May 17]. Available from: https://www.census.gov/quickfacts/fact/table/US/PST045218

37. Benthin A, Severson H, Slovic P. A psychometric study of adolescent risk perception.

38. John OP, Donahue EM, Kentle RL. The big five inventory—versions 4a and 54. 1991.

39. John OP, Naumann LP, Soto CJ. Paradigm shift to the integrative big five trait taxonomy. Handbook of personality: Theory and research. 2008 Aug 5;3(2):114–58.

40. Rammstedt B, John OP. Measuring personality in one minute or less: A 10-item short version of the Big Five Inventory in English and German. Journal of research in Personality. 2007 Feb 1;41(1):203–12.

41. Davis MH. Measuring individual differences in empathy: Evidence for a multidimensional approach. Journal of personality and social psychology. 1983 Jan;44(1): 113.

42. Roets, A., & Van Hiel, A. (2011). Item selection and validation of a brief, 15-item version of the Need for Closure Scale. Personality and Individual Differences, 50(1), 90–94.

43. Carver CS, White TL. Behavioral inhibition, behavioral activation, and affective responses to impending reward and punishment: the BIS/BAS scales. Journal of personality and social psychology. 1994 Aug;67(2): 319.

44. Gray JA. A critique of Eysenck’s theory of personality. InA model for personality 1981 (pp. 246–276). Springer, Berlin, Heidelberg.

45. Gray JA. The psychology of fear and stress. Cambridge University Press. New York. 1987.

46. Budescu, D.V. (1993). Dominance analysis: A new approach to the problem of relative importance of predictors in multiple regression, Psychological bulletin 114 (3), 542.

47. Norušis MJ. IBM SPSS Statistics 19 Statistical Procedures Companion. Prentice Hall; 2011. 646 p.

48. Li CH. Confirmatory factor analysis with ordinal data: Comparing robust maximum likelihood and diagonally weighted least squares. Behavior research methods. 2016 Sep 1;48(3):936–49.

49. Zygmont C, Smith MR. Robust factor analysis in the presence of normality violations, missing data, and outliers: Empirical questions and possible solutions. The Quantitative Methods for Psychology. 2014;10(1):40–55.

50. Baumeister RF, Scher SJ. Self-defeating behavior patterns among normal individuals: review and analysis of common self-destructive tendencies. Psychological bulletin. 1988 Jul;104(1): 3.

51. Farmer PE, Nizeye B, Stulac S, Keshavjee S. Structural violence and clinical medicine. PLoS medicine. 2006 Oct;3(10).

52. Stanger-Hall KF, Hall DW. Abstinence-only education and teen pregnancy rates: why we need comprehensive sex education in the U.S.PLoS ONE. 2011;6(10): e24658.

53. Arkowitz SO, Lilienfeld H. Why “Just Say No” Doesn’t Work [Internet]. Scientific American. 2014 [cited 2020 May 12]. Available from: https://www.scientificamerican.com/article/why-just-say-no-doesnt-work/

